# DNA methylation alterations in *MIR10B*, *MIR21*, *MIR100*, *MIR127* and *MIR143* genes associated with advanced carotid artery atherosclerosis: a targeted bisulfite sequencing study of vascular tissues and peripheral blood

**DOI:** 10.64898/2026.08.03.26359571

**Authors:** Iuliia A. Koroleva, Alexey A. Zarubin, Anton V. Markov, Alexey A. Sleptcov, Mikhail S. Kuznetsov, Boris N. Kozlov, Elvira F. Muslimova, Sergey A. Afanasiev, Nadezhda P. Babushkina, Elena Yu. Bragina, Irina A. Goncharova, Maria V. Golubenko, Aksana N. Kucher, Maria S. Nazarenko

**Affiliations:** Research Institute of Medical Genetics, Tomsk National Research Medical Center, Tomsk, Russia; Cardiology Research Institute, Tomsk National Research Medical Center, Tomsk, Russia

## Abstract

DNA methylation is a key epigenetic mechanism regulating the expression of genes involved in numerous developmental and pathological processes. However, the contribution of DNA methylation of microRNA genes to atherosclerosis remains poorly understood.

In this study, we profiled DNA methylation patterns of both the regulatory elements and gene bodies of five microRNA genes (*MIR10B*, *MIR21*, *MIR100*, *MIR127,* and *MIR143*) in vascular tissues and paired peripheral blood cells (PBC) of 92 patients with advanced carotid atherosclerosis and 32 PBC of control participants by targeted bisulfite sequencing. We identified distinct tissue-specific DNA methylation patterns for all five microRNA genes in patients with advanced carotid atherosclerosis. The regulatory regions of *MIR10B*, *MIR127*, and *MIR100* were moderately hypomethylated in carotid atherosclerotic plaques compared with intact vascular tissues.

We further integrated our findings with lab-internal and publicly available epigenome-wide methylation datasets and evaluated the influence of vascular and blood cell composition using computational deconvolution approaches. After adjustment for cellular heterogeneity in vascular tissues, DNA methylation at CpG sites in *MIR100* and *MIR127* remained independently associated with atherosclerosis. Increased DNA methylation at a single CpG site (chr11:122025143, GRCh37/hg19) located within the *MIR100* E-box region was associated with metabolic syndrome. Moreover, DNA methylation levels of *MIR10B*, *MIR21*, and *MIR127* in atherosclerotic plaques were linked with indicators of histological instability and history of acute cerebrovascular events.

In peripheral blood, we observed moderate hypomethylation of the regulatory regions of *MIR10B*, *MIR21*, and the *MIR100* E-box region in patients compared with the control group. However, only the *MIR10B* remained robust against blood cell composition. In blood, *MIR10B* and *MIR143* methylation correlated with lipid metabolism and carotid stenosis, while the *MIR21* CpG island showed strong blood-plaque concordance, confirming its potential as a surrogate biomarker.

Overall, advanced carotid atherosclerosis is characterized by specific tissue-altering DNA methylation patterns of microRNA genes, where alterations mainly occur in the regulatory regions, predominantly featuring hypomethylation. The results underscore the complex, cell-and tissue-specific nature of DNA methylation of microRNA genes in both the regulatory elements and gene bodies in vascular tissue and blood, highlighting the critical need to decipher these intricate epigenetic landscapes to identify reliable, robust biomarkers for assessing plaque instability and cardiovascular risk.

## Background

Atherosclerosis (AS) is a complex lesion of arteries characterized by chronic inflammation, lipid accumulation, and vascular wall remodeling. In advanced stages, rupture or erosion of atherosclerotic plaques triggers platelet adhesion, thrombus formation, arterial occlusion and acute ischemic events, including coronary artery disease, myocardial infarction, and ischemic stroke, which remain the leading cause of mortality worldwide [Volný O. et al., 2015; Li Z. et al., 2023; Euler G., Parahuleva M., 2025].

The pathogenesis of AS is regulated by multiple classes of RNA molecules, including microRNAs (miRNAs), which are small single-stranded non-coding RNAs, typically 16-27 nucleotides in length, that regulate gene expression at the post-transcriptional level [Fang Z. et al., 2013; Vartak T. et al., 2022; Wang J. et al., 2025]. Approximately 2,500 miRNAs have been identified in the human genome, of which nearly 150 microRNAs play a critical role in the cardiovascular system [Cordeiro M.A. et al., 2025].

MicroRNAs dichotomously regulate a wide range of biological processes in AS including initiation (endothelial senescence and inflammation), progression (SMC proliferation and migration, increased necrosis of lipid core, angiogenesis), and endpoint (unstable plaque rupture) [Wang J. et al., 2025]. According to their effects on atherosclerosis progression, microRNAs can be divided into proatherogenic, antiatherogenic or ambigual groups [Volný O. et al., 2015; Euler G., Parahuleva M., 2025; Wang J. et al., 2025].

Numerous studies have investigated alterations in microRNA expression in human carotid atherosclerotic lesions using various groups of comparison patient cohorts and different experimental approaches [Cipollone F. et al. 2011; Lovren F. et al., 2012; Nazari-Jahantigh M. et al., 2012; Cao J. et al., 2014; Bazan H.A. et al., 2015; Maitrias P. et al., 2015; Markus B. et al, 2016; Jin H. et al., 2018; Katano H. et al., 2018; Wang H. et al., 2019; Collura S. et al., 2021; González-López P. et al., 2022; Nazarenko M.S. et al., 2022; Deng Y. et al., 2023]. Although the reported findings vary considerably and only partially overlap, by the end of 2024, more than two independent studies had demonstrated altered expression of miR-21, miR-143/-145, miR-100, and miR-127 during the development and destabilization of carotid atherosclerotic plaques [Koroleva I.A. et al., 2017; Jin H. et al., 2018; Katano H. et al., 2018; Wang H. et al., 2019; González-López P. et al., 2022; Nazarenko M.S. et al., 2022].

MicroRNA expression is regulated by multiple mechanisms, including epigenetic modifications such as DNA methylation of regulatory genomic regions. Several types of regulatory elements may contribute to methylation-dependent regulation of the microRNA genes, including: (1) promoter regions enriched in CpG sites capable to methylation; (2) CpG islands located in the proximal promoter regions of microRNA gene; (3) CpG islands upstream of transcription start site (TSS) of microRNA cluster; (4) CpG island located in the promoter of the host gene of intronic microRNA; (5) distant enhancers that can regulate microRNA expression [Morales S. et al., 2017].

Most studies investigating DNA methylation of microRNA regulatory regions have focused on cancer. These studies have demonstrated that hypermethylation of promoter-associated CpG islands is commonly associated with reduced microRNA expression, whereas promoter or enhancer hypomethylation may result in transcriptional activation [Baer C. et al., 2013; Chhabra R., 2014; Bell R.E. et al., 2016; Morales S. et al., 2017; Ali Syeda Z. et al., 2020; Saviana M. et al., 2023]. In contrast, the contribution of DNA methylation of microRNA genes to other common diseases, including atherosclerosis, remains poorly understood [Ek W.E. et al., 2016; Sharma A.R. et al., 2020; Cui C. et al., 2024].

Epigenome-wide association studies (EWAS) typically examine DNA methylation at CpG sites within an intergenic region or inside protein-coding genes in peripheral blood cells from patients with different manifestations of atherosclerosis [Istas G. et al., 2017; Liu Y. et al., 2017; Soriano-Tárraga C. et al., 2018; Sharma A.R. et al., 2020; Ingold M.et al., 2026]. Comparatively few studies have investigated DNA methylation changes in vascular tissues or atherosclerotic plaques [Castillo-Díaz S.A. et al., 2010; Fernandez A.F. et al., 2011; Zaina S. et al., 2014; Aavik E. et al., 2015; Nazarenko M.S. et al., 2015; Zaina S. et al., 2015; Li J. et al., 2021]. Previous studies comparing advanced atherosclerotic and intact vascular tissues have reported both decreased DNA methylation in genomic regions encompassing *MIR10B* and *MIR127* [Aavik E. et al., 2015; Nazarenko M.S. et al., 2015] and increased DNA methylation within the *MIR143/MIR145* cluster [Ehrlich K.C. et al., 2019].

To date, no studies have comprehensively examined DNA methylation level of regulatory elements of microRNA genes in carotid atherosclerotic plaques. Because DNA methylation represents a potential mechanism regulating microRNA expression, characterization of DNA methylation patterns within both regulatory regions and gene bodies of microRNA genes associated with plaque instability is warranted in carotid atherosclerotic plaques and peripheral blood leukocytes from patients with carotid atherosclerosis. Identifying disease-associated DNA methylation patterns in vascular tissues and blood may improve our understanding of the epigenetic mechanisms underlying advanced atherosclerosis and facilitate the development of novel biomarkers for predicting plaque instability and atherosclerosis complication.

## Methods

### 1. Study participants and sample collection

This study included 92 patients with advanced carotid atherosclerosis (70% men, median age 64 years) and 32 control participants without symptoms of cerebrovascular disease (60% men, median age 67 years) (Supplementary Methods - Table SM1). All participants underwent carotid ultrasonography. The patient group comprised individuals with severe carotid stenosis (>65%), an indication for carotid endarterectomy. Control participants had early-stage carotid atherosclerosis without hemodynamically significant changes: the degree of stenosis was 28 [24; 45]% (M [Q1; Q3]), and intima-media complex thickness was 0.8 [0.7; 0.9] mm. Peripheral blood samples were collected from each control participant (PBC). The study was conducted in accordance with the Declaration of Helsinki and approved by the Local Biomedical Ethics Committee at the Research Institute of Medical Genetics, Tomsk National Research Medical Center (protocol #4, 25 November 2019).

Peripheral blood samples were collected from each patient before carotid endarterectomy (PBP; n=92). During surgery, carotid atherosclerotic plaque samples (ASP; n=92), macroscopically intact adjacent carotid artery tissue (ICA; n=34), and great saphenous vein samples (GSV; n=23) were obtained. Each ASP sample was divided into 3 equal fragments with similar macroscopic characteristics. One fragment was used for histological verification of the ASP type according to the American Heart Association (AHA) system [Stary H.C. et al. 1995], assessment of plaque stability, and evaluation of pathomorphological characteristics (Supplementary Methods). A second fragment was used for DNA extraction, sodium bisulfite treatment, and DNA methylation analysis. The third fragment was placed back into a tube containing a reagent for RNA stabilization and sample storage in RNALater (Dia-m, Russia) for subsequent analysis.

### 2. DNA methylation analysis in vascular tissues and peripheral blood

To identify the genomic locations of regulatory elements of selected *MIR* genes and miRNA host genes, we used FANTOM5 (http://fantom.gsc.riken.jp/5/) [de Rie D. et al., 2017] and TRmir (http://bio.liclab.net/trmir/index.html) databases [Gao Y. et al., 2022], together with publications indexed in HMDD v3.2: the Human miRNA Disease Database (http://www.cuilab.cn/hmdd) [Cui C. et al., 2024] and NCBI: National Center for Biotechnology Information (https://www.ncbi.nlm.nih.gov).

DNA methylation at CpG sites within the target regions was quantified using **Bisulfite Amplicon Sequencing (BSAS)** performed on the MiSeq platform (Illumina, USA) using massive parallel sequencing [Masser D.R. et al., 2015]. All the BSAS stages, as well as the target region selection procedure, and genomic coordinates of the analyzed CpG sites are described in Supplementary Methods.

To examine methylation at CpG sites associated with miRNA genes using microarray data, we analyzed lab-internal EWAS dataset of DNA methylation and microRNA expression in ASP samples (unpublished data). A total of 16 ASP samples were selected according to histological stability and divided into two equal groups: 8 type III-V plaques (preatheroma and stable lesions) and 8 type VI plaques (complicated lesions), based on the AHA classification (Supplementary Methods - Table SM2). The corresponding patients comprised 10 men and 6 women, with a median age of 65 years (Supplementary Methods - Table SM3). DNA methylation was profiled using the Illumina Infinium MethylationEPIC BeadChip microarray.

### 3. Data processing and statistical analysis

BSAS data were processed using established bioinformatic methods and pipelines (Supplementary Methods). For each CpG site, the methylation level was expressed as beta (β) value, calculated as the percentage of reads corresponding to methylated cytosine among all cytosine reads at a specific genomic position.

For lab-internal EWAS, microarray signal intensities were scanned using an iScan® System scanner (Illumina, USA) and recorded as images and raw IDAT files. These data were used to analyze the level of DNA methylation using Bioconductor packages [Aryee M.J. et al., 2014] in the R, free software environment for statistical computing and graphics [R Core Team, 2025]. The data on the CpG sites of microRNA genes provided in the annotation of the microarray manufacturer (Illumina, USA) have been updated in accordance with the human genome version GRCh37/hg19 and supplemented with information on miRNA gene regulatory elements from the TRmir database (http://bio.liclab.net/trmir/index.html) [Gao Y. et al., 2022].

Statistical analyses were performed in R (version 4.5.2 and earlier versions) using standard packages (R Core Team, 2025). CpG-site methylation levels are reported as M [Q1; Q3], where M denotes the median and Q1 and Q3 denote the first (25th percentile) and third (75th percentile) quartiles, respectively.

Differences in DNA methylation between study groups were assessed using nonparametric tests. The Wilcoxon test was used for paired comparisons of different tissue types obtained from the same patients, whereas the Mann-Whitney U test was used for independent comparisons of peripheral blood leukocytes from patients and controls. Correlations were assessed using Spearman’s rank correlation, and coefficient magnitudes were interpreted according to Schober et al. (2018) [Schober et al., 2018]. A p value < 0.05 was considered statistically significant. The Benjamini-Hochberg procedure was used to control the false discovery rate (FDR) in multiple hypothesis testing [Benjamini Y., Hochberg Y., 1995]. FDR correction was applied separately within each table in Results.

To compare our findings with external data on microRNA gene methylation in vascular tissues and blood cells we selected five publicly available DNA methylation datasets from the Gene Expression Omnibus (GEO) public data repository (https://www.ncbi.nlm.nih.gov/geo): GSE46394 [Zaina S. et al., 2014], GSE66500 [Zaina S. et al., 2015], GSE149759 [Li J. et al., 2021], GSE69138 [Soriano-Tárraga C. et al., 2018], and GSE246337 [Moqri M. et al., 2026] (Supplementary Table 11).

### 4. Estimation of cell-type fractions in vascular tissues and peripheral blood

Cell-type fractions in vascular tissues and peripheral blood were estimated using two categories of DNA methylation (DNAm) datasets: external microarray datasets (GSE46394, GSE66500, GSE149759, and GSE246337) and the lab-internal EWAS dataset described above. External datasets were downloaded from GEO as normalized beta values. For vascular tissues, original DNAm reference matrices (Supplementary Table 12) and the HEpiDISH algorithm [Teschendorff A.E. et al. 2017; Zheng S.C. et al. 2018] were used. Performance of the vascular cell-type estimation was evaluated using 28 miRNA-associated CpG sites represented on Illumina methylation microarrays. We identified CpG sites specific to smooth muscle cells (SMCs) and immune cells (ICs) and created the model for targeted bisulfite sequencing data of miRNA genes to determine whether the disease status or proportion of vascular cells contribute to variations in microRNA gene methylation. For peripheral blood we chose one DNAm reference matrix designed for adult whole blood in EpiDISH algorithm [Teschendorff A.E. et al. 2017]. Blood cell-type estimation in the external EWAS dataset GSE246337 was evaluated using the same 28 miRNA-associated CpG sites on Illumina methylation microarrays.

## Results

### 1. Tissue specificity of DNA methylation level of MIR10B, MIR21, MIR100, MIR127 and MIR143 gene regions between blood and vascular tissues

We analyzed DNA methylation in eight regions related to five microRNA genes:

1) The CpG island overlaying *MIR10B* gene body; 23 CpG sites (hereafter referred to as ***MIR10B* region**);
2) The literature-defined putative promoter of *MIR21* gene; 9 CpG sites (***MIR21* putative promoter**);
3) The ***MIR21* CpG island** described by Iorio M.V. et al. (2007) [Iorio M.V. et al., 2007]; 16 CpG sites;
4) The ***MIR21* gene body**; 5 CpG sites;
5) The E-box region containing the transcription factor ZEB1 binding site and representing the putative regulatory region of *MIR100* gene [Chen D. et al., 2014]; 7 CpG sites (***MIR100* E-box region**);
6) The CpG island overlapping with *MIR127* gene body; 40 CpG sites (***MIR127* region**);
7) The ***MIR143HG* promoter**, representing the regulatory region of *MIR143* gene; 10 CpG sites;
8) The ***MIR143* gene body**; 5 CpG sites.

For the intergenic miRNA genes *MIR10B* and *MIR127*, the selected regions comprised CpG islands that fully or partially overlapped the gene bodies. For the intragenic genes *MIR21* and *MIR143*, putative regulatory regions and gene bodies were analyzed; the *MIR21* CpG island was also analyzed.

Across all eight regions, median DNA methylation patterns were tissue-specific. Methylation profiles were highly similar between peripheral blood leukocytes from patients (PBP) and controls (PBC) but differed between blood leukocytes and vascular tissues, including carotid atherosclerotic plaques (ASP), macroscopically intact adjacent carotid artery tissue (ICA), and great saphenous veins tissue (GSV) (Figures 1,2; Supplementary Tables 01-09).

**Figure 1:**
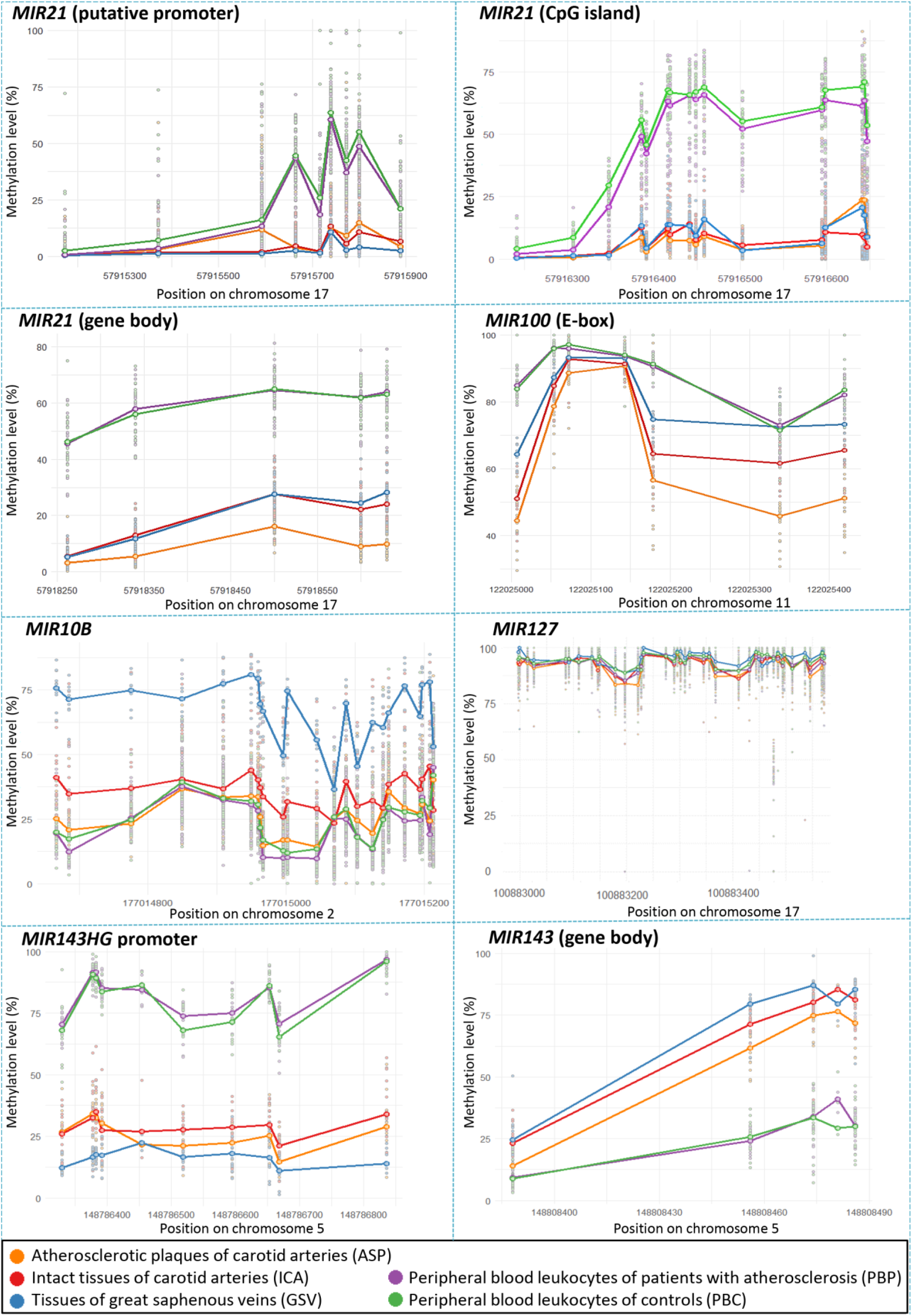
DNA methylation profiles across the investigated microRNA gene regions. Genomic coordinates are given according to the GRCh37/hg19 genome assembly. Each point represents an individual CpG methylation value, and lines connect the median methylation levels within each tissue.

**Figure 2:**
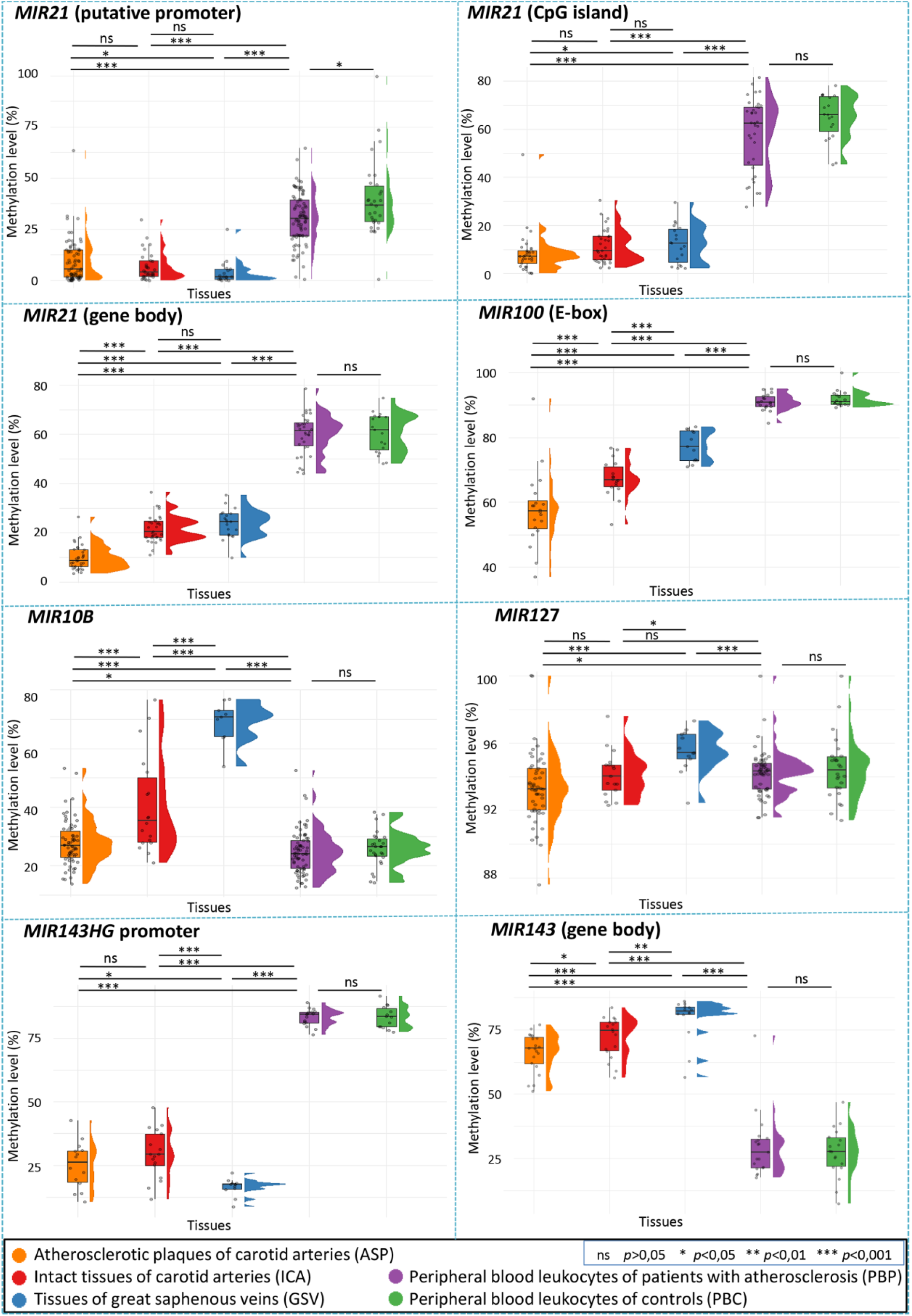
Comparison of median DNA methylation across the investigated microRNA gene regions. Each point represents the median methylation level across all CpG sites within the corresponding region for an individual sample.

In the *MIR127* region, DNA methylation levels were generally high (80–100%) across all tissues (in Figure 1, the upward shift of the plots for *MIR127* should be noted). However, a distinct cluster of outlying samples with approximately 50% methylation was consistently observed at CpG site chr14:101349764 (here and further, GRCh37/hg19 coordinates). This CpG overlaps with the single nucleotide variant rs34163305:C>T, for which the minor T allele has a frequency of 0.24 in the European population according to gnomAD v.4.1.0 database [https://gnomad.broadinstitute.org/variant/14-100883427-C-T]. Because the T allele abolishes the CpG dinucleotide, methylation cannot occur in TT homozygotes, whereas values of approximately 50% are expected in heterozygotes.

Median methylation was higher in leukocytes (PBP and PBC) than in vascular tissues (ASP, ICA, and GSV) at all three ***MIR21*** regions: the **putative promoter** (min-max: 30.3%-36.8% vs. 1.7-5.7%), **CpG island** (min-max: 62.4-66.1% vs. 7.2-12.7%) and **gene body** (min-max: 61.4-61.8% vs. 8.7-24.5%). Similarly elevated methylation in leukocytes was observed in the ***MIR100* E-box region** (min-max: 90.9-91.0% vs. 57.3-77.2%) and ***MIR143HG* promoter** (min-max: 83.5-84.4% vs. 17.4-29.3%) (Figure 2; Supplementary Tables 01-09).

By contrast, median methylation of the ***MIR143* gene body** was lower in PBP and PBC than in vascular tissues (min-max: 27.5-27.7% vs. 67.8-82.2%, *p*<0.001). Median methylation of the ***MIR10B*** region was markedly lower in PBP and PBC than in GSV (min-max: 24.0-26.5% vs. 70.8%). Likewise, median methylation across the region was lower in leukocytes than in carotid tissues (ASP and ICA; min-max: 24.0-26.5% vs. 26.9-35.4%) (Figure 2). However, these differences at individual CpG sites were small or not statistically significant (Figure 1; Supplementary Tables 01-09).

The ***MIR127* region** including CpG island was highly methylated (>80%) in all tissues except at the chr14:101349764 described above (Figure 1; Supplementary Table 07). Similar to *MIR10B* region, the highest median methylation was detected in GSV (Figure 2; Supplementary Table 07). Although several comparisons reached statistical significance, the magnitude of these differences was small, not exceeding 2.2%.

### 2. Methylation of microRNA genes in vascular tissues of patients with carotid artery atherosclerosis

#### 2.1 CpG islands of MIR10B and MIR127 genes, gene bodies of MIR21 and MIR143, and MIR100 regulatory region are moderately hypomethylated in carotid atherosclerotic lesions

Compared with ICA, ASP demonstrated significantly lower median DNA methylation (Figure 2; Supplementary Tables 01-09) in the ***MIR10B*** region (by 8.5%, *p*<0.001), ***MIR21* gene body** (by 11.7%, *p*<0.001), ***MIR100* E-box region** (by 9.7%, *p*<0.001) and ***MIR143* gene body** (by 6.9%, *p*=0.019). Median methylation in these regions was also lower in ASP than in GSV; the same pattern was observed for ***MIR127*** region (Figure 2; Supplementary Tables 01-09).

To validate these findings, our BSAS results were compared with lab-internal EWAS dataset and publicly available EWAS datasets from GEO (the genomic correspondence between BSAS target regions and Illumina methylation microarrays probes is summarized in Supplementary Table 10. Notably, the *MIR100* E-box region does not contain CpG sites annotated on Illumina microarray platform).

Methylation patterns for *MIR10B*, *MIR21* and *MIR143* were highly consistent across independent studies, analytical platforms, and vascular tissues, except for the *MIR21* gene body in the internal mammary artery (IMA) samples from GSE149759 (Figure 3). In contrast, methylation of cg21242144 within the *MIR143* gene body differed substantially between BSAS and microarray datasets.

**Figure 3:**
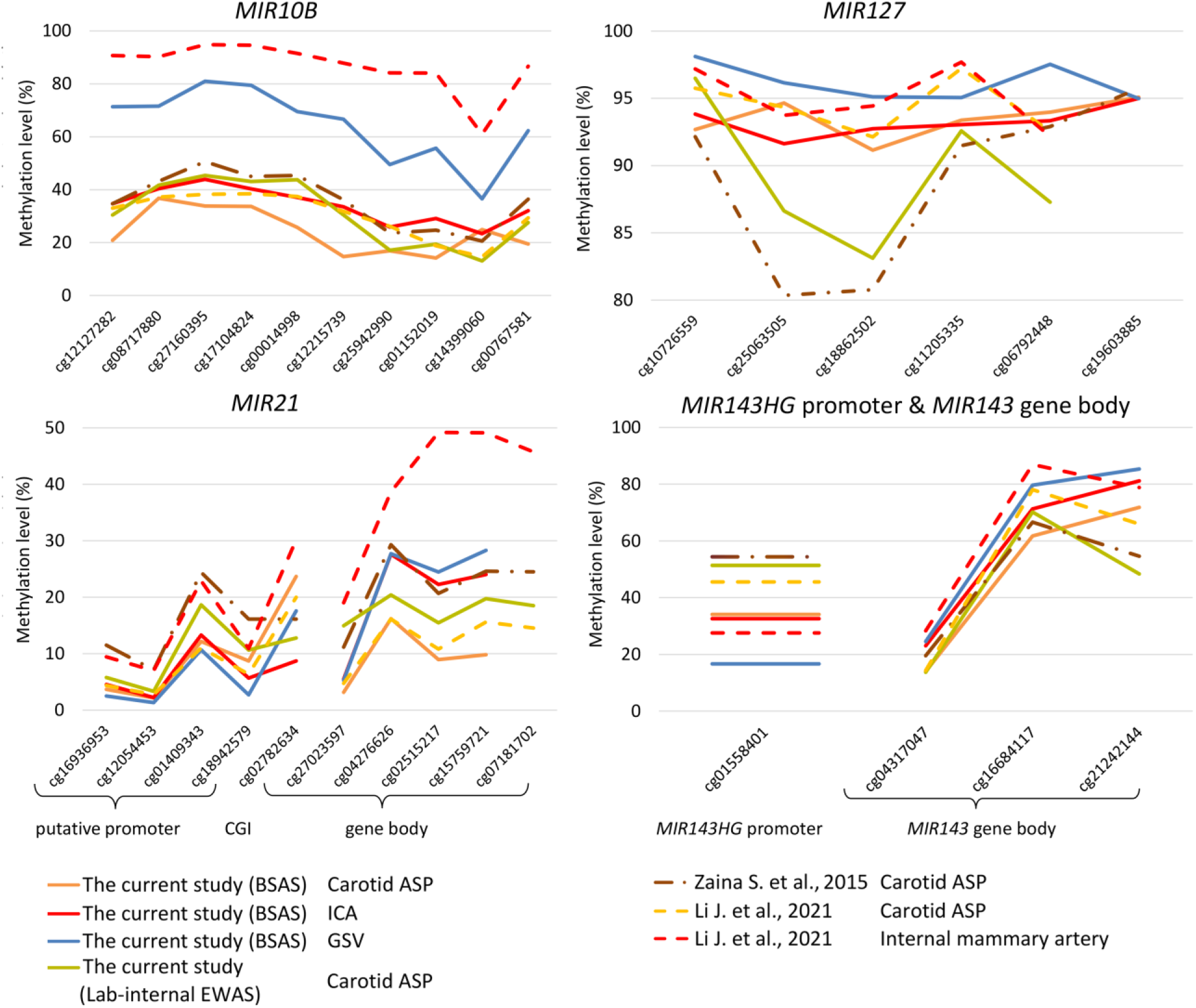
CpG methylation patterns in different blood vessels and carotid plaques measured by bisulfite amplicon sequencing (BSAS) and Illumina methylation microarrays technology (external and lab-internal EWAS). ASP, carotid atherosclerotic plaques; ICA, intact carotid artery; GSV, great saphenous veins.

The methylation profile of ASP samples in lab-internal EWAS was similar to that reported by S. Zaina et al. (2015), whereas greater discrepancies were observed relative to the dataset published by J. Li et al. (2021). Because all three studies used Illumina methylation arrays, these inconsistencies are likely attributable to biological rather than technical factors, including differences in patient age, ethnicity, vascular cell composition, and disease characteristics.

Across all CpG sites in the ***MIR10B*** region, ***MIR21* gene body** and ***MIR143* gene body**, methylation was lower in ASP than in IMA or GSV in BSAS and in microarray datasets (Figure 3) [Zaina S. et al., 2015; Li J. et al., 2021; lab-internal EWAS (unpublished data)]. In contrast, ***MIR127*** methylation was uniformly high (>80%) across all vascular tissues, without a clear disease-specific methylation pattern. Nevertheless, analysis of BSAS data separately suggested lower methylation in ASP than in GSV (Figure 3).

Unlike the other analyzed regions, the ***MIR143HG* promoter**, represented by single cg01558401 site, exhibited higher DNA methylation in ASP than in unaffected vessels (min-max: 34.2%-54.4% vs. 16.7-32.6%) (Figure 3) [Zaina S. et al., 2015; Li J. et al., 2021; lab-internal EWAS (unpublished data)].

#### 2.2. Variation of MIR100 and MIR127 gene methylation of vascular tissue is due to atherosclerosis, but not cellular heterogeneity

Cellular heterogeneity is one of the most significant confounding factors in DNA methylation research because blood vessels contain variable proportions of stromal and immune cell populations, each characterized by distinct DNA methylation profiles. Consequently, observed methylation differences reflect shifts in cellular composition rather than providing accurate insights into the trait being investigated [Eipel M. et al. 2016; Berg F. et al., 2025].

To account for this source of variation, cell-type proportions were estimated using 102 vascular samples data derived from combined external EWAS datasets (GSE149759, GSE46394, GSE66500) and lab-internal EWAS (Supplementary Table 11). The HEpiDISH algorithm [Teschendorff A.E. et al. 2017; Zheng S.C. et al. 2018] was applied using original DNA methylation reference matrices for vascular cells (Supplementary Table 12), enabling estimation of ten major vascular cell populations, including vascular endothelial cells (VECs), smooth muscle cells (SMCs), fibroblasts (FIB) and immune cells (ICs): B-cells, NK, CD4T-cells, CD8T-cells, monocytes (Mono), neutrophils (Neutro), eosinophils (Eosino) (Supplementary Table 13).

Because SMCs, VECs and immune cells represent the principal cellular populations that change during plaque progression, these variables were selected for adjustment. We subsequently examined associations between DNA methylation at the 28 analyzed CpG sites of microRNA genes annotated on the Illumina platform (Supplementary Table 10) and the estimated proportions of VECs, SMCs, and IC using Weighted Gene Co-expression Network Analysis (WGCNA) [Langfelder P., Horvath S., 2012].

These deconvolution data demonstrate a clear cell-type dichotomy, where ***MIR10B***, ***MIR127*** (except cg25063505), and ***MIR143* gene body** hypermethylation characterize SMCs, whereas elevated methylation of ***MIR21*** and ***MIR143HG* promoter** aligns with IC accumulation in vascular wall.

The strongest positive association was observed between methylation of cg01558401 within the ***MIR143HG* promoter** and the estimated ICs proportion (*rho* = 0.96, *p* < 0.001) (Figure 4). Two CpG sites within the ***MIR143* gene body** (cg16684117 and cg21242144) showed similarly strong positive correlations with SMCs percentage (*rho* = 0.92 and 0.91 respectively) (Figure 4).

**Figure 4.**
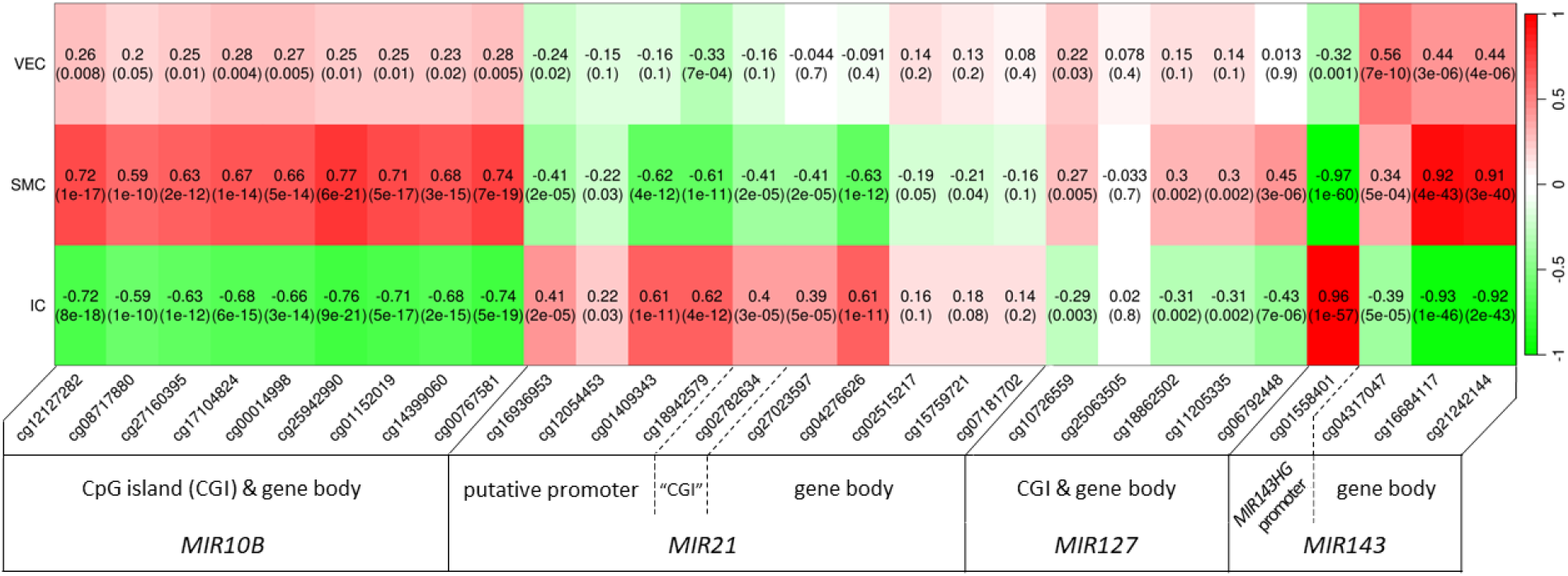
Correlation heatmap between DNA methylation of analyzed CpG sites and estimated proportions of vascular endothelial cells (VECs), smooth muscle cells (SMCs) and immune cells (ICs).

Based on these findings, cg01558401 (in ***MIR143HG* promoter**) and cg21242144 (in ***MIR143* gene body**) were selected for further analysis because they exhibited the strongest associations with estimated ICs and SMCs fractions and were also covered by BSAS.

As expected, the opposite relationships of these CpG sites with ICs or SMCs abundance resulted in a strong inverse correlation between their methylation levels. This relationship was observed consistently in the combined EWAS datasets (*rho* =-0.801, *p* < 0.001) (Figure 5A) and independently confirmed in the BSAS cohort (*rho* =-0.784, *p* < 0.001) (Figure 5B).

**Figure 5:**
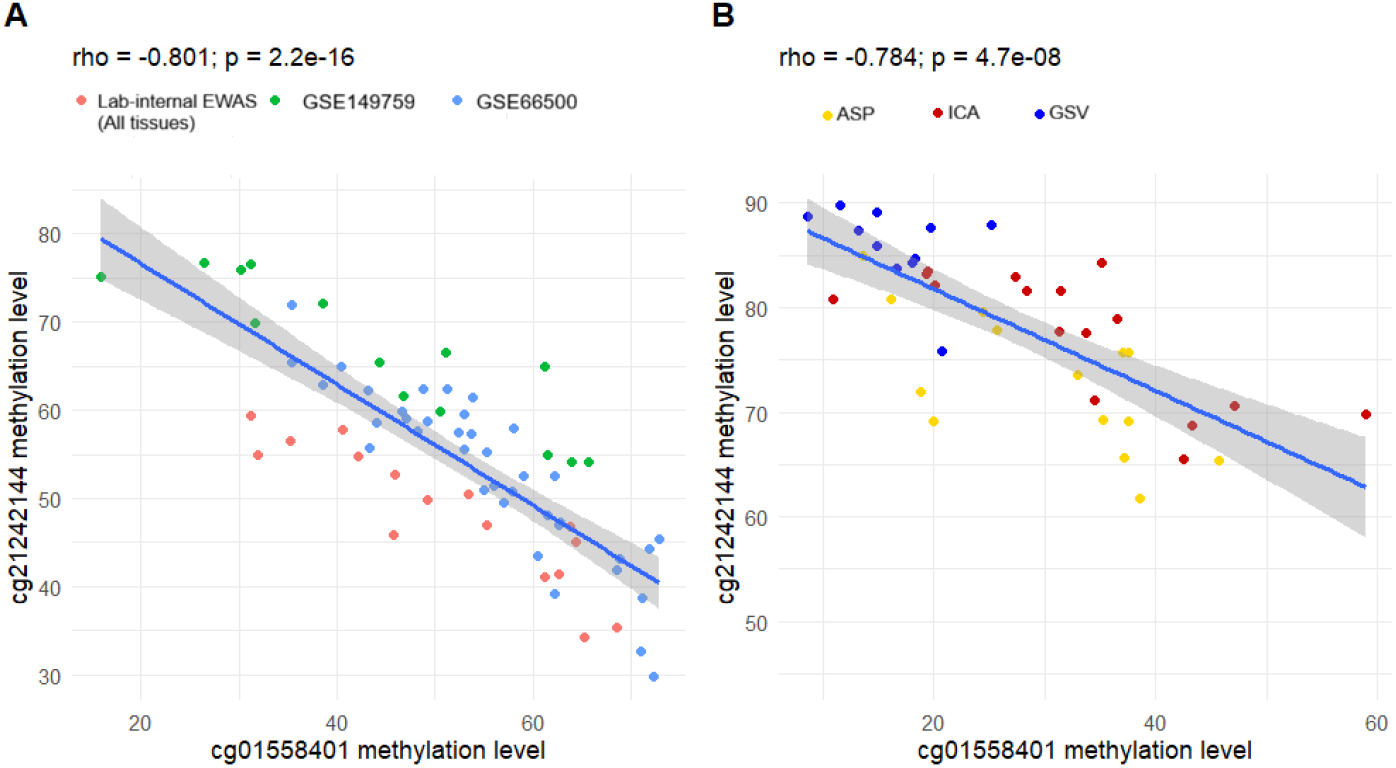
Correlation between DNA methylation of cg01558401 (in ***MIR143HG* promoter**) and cg21242144 (in ***MIR143* gene body**) in the combined EWAS datasets (A) and BSAS cohort (B).

To exclude age as a potential confounder, we examined the relationship between methylation at these CpG sites and patient age. No significant associations were detected (cg01558401: *rho* =-0.091, *p* =0.604; cg21242144: *rho* = 0.113, *p* =0.519), indicating that methylation of two target CpG sites remains stable within the studied age range (58-70 years).

Then we generated linear models to estimate the proportion of SMCs/ICs based on BSAS methylation values of cg01558401 and cg21242144. The intercepts and coefficients of these models were extracted to formulate a general-purpose estimation of the proportion of SMCs and ICs.

**Estimated % of Smooth Muscle Cells** = 0.6266 – 0.9531 × cg01558401 methylation + 0.5454 × cg21242144 methylation.

**Estimated % of Immune Cells** = 0.3856 + 0.9102 × cg01558401 methylation – 0.6877 × cg21242144 methylation.

Finally, we fitted regression models to the targeted BSAS data to evaluate associations of CpG methylation with carotid artery state (ICA or ASP) and estimated SMC or IC proportion (Table 1) and to determine whether DNA methylation differences reflected changes in vascular cell composition or disease-associated functional changes in carotid artery cells.

**Table 1.**
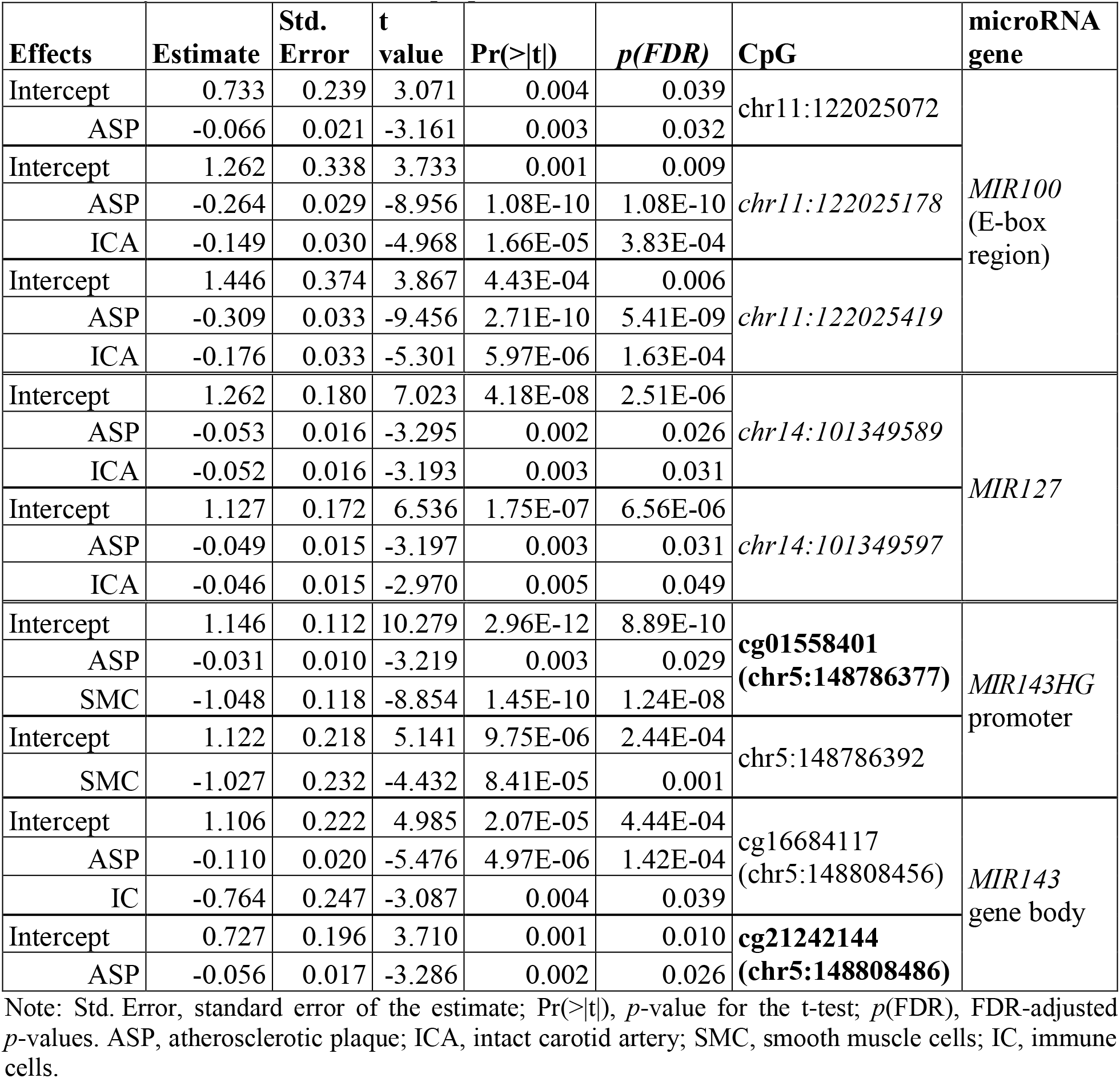
Regression analysis of CpG sites in microRNA genes methylation levels in relation to carotid artery state and the estimated proportion of vascular cells.

Models were adjusted for all covariates. CpG sites used to generate the model for estimation are shown in **bold**. CpG sites influenced by *both* carotid atherosclerosis (ASP) and normal artery state (ICA) – that is, rather by functional changes in carotid artery cells – but not by estimated proportion of smooth muscle cells (SMCs) or immune cells (ICs) are shown in *italics*.

In these models, four CpG sites, two in the *MIR100* E-box region and two in the *MIR127* region, remained significantly associated with carotid atherosclerosis after adjustment for estimated SMC or IC proportions. Thus, methylation at these sites was associated with carotid artery state independently of the included cell-composition estimates, indicating that their methylation changes most likely reflect disease-related functional alterations in vascular cells rather than differences in cellular heterogeneity.

#### 2.3. DNA methylation levels of the MIR10B, MIR21, and MIR127 genes in carotid atherosclerotic plaque are associated with indicators of histological plaque instability and acute cerebrovascular events

We investigated the associations between the DNA methylation levels across all analyzed microRNA gene regions in carotid ASP and both qualitative and quantitative indicators of histological plaque instability.

Within the ***MIR21* putative promoter**, methylation levels of three CpG sites (chr17:57915593, cg12054453 and cg18942579), as well as the median methylation level across the entire region, were significantly lower in ASPs with intraplaque hemorrhage in the plaque shoulder than in ASP without hemorrhages (Table 2). Similarly, five of the nine analyzed CpG sites, including the three sites mentioned above, together with the median methylation level across the region, exhibited significantly reduced methylation in ASPs containing foam cells (Table 2).

**Table 2.**
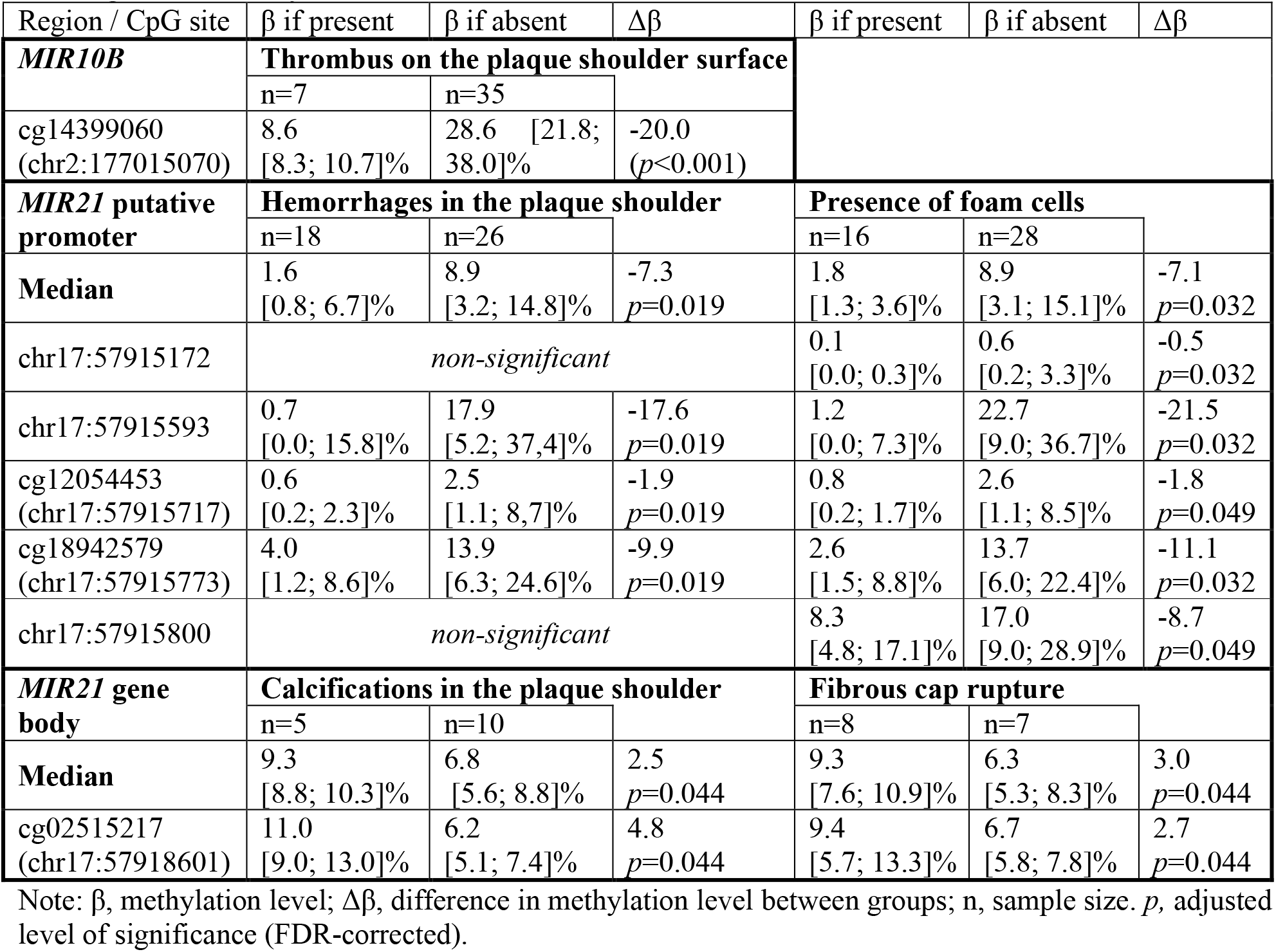
Statistically significant changes of methylation level in CpG sites and entire microRNA genes regions depending on individual <u>qualitative</u> indicators of the atherosclerotic plaque histological instability.

| Region / CpG site | $\beta$ if present | $\beta$ if absent | $\Delta\beta$ | $\beta$ if present | $\beta$ if absent | $\Delta\beta$ |
| --- | --- | --- | --- | --- | --- | --- |
| ***MIR10B*** | **Thrombus on the plaque shoulder surface** |  |  |  |  |  |
|  | n=7 | n=35 |  |  |  |  |
| cg14399060<br>(chr2:177015070) | 8.6<br>[8.3; 10.7]% | 28.6 [21.8;<br>38.0]% | -20.0<br>($p<0.001$) | | | |
| ***MIR21* putative promoter** | **Hemorrhages in the plaque shoulder** |  |  | **Presence of foam cells** |  |  |
|  | n=18 | n=26 |  | n=16 | n=28 |  |
| **Median** | 1.6<br>[0.8; 6.7]% | 8.9<br>[3.2; 14.8]% | -7.3<br>$p=0.019$ | 1.8<br>[1.3; 3.6]% | 8.9<br>[3.1; 15.1]% | -7.1<br>$p=0.032$ |
| chr17:57915172 | *non-significant* | | | 0.1<br>[0.0; 0.3]% | 0.6<br>[0.2; 3.3]% | -0.5<br>$p=0.032$ |
| chr17:57915593 | 0.7<br>[0.0; 15.8]% | 17.9<br>[5.2; 37.4]% | -17.6<br>$p=0.019$ | 1.2<br>[0.0; 7.3]% | 22.7<br>[9.0; 36.7]% | -21.5<br>$p=0.032$ |
| cg12054453<br>(chr17:57915717) | 0.6<br>[0.2; 2.3]% | 2.5<br>[1.1; 8.7]% | -1.9<br>$p=0.019$ | 0.8<br>[0.2; 1.7]% | 2.6<br>[1.1; 8.5]% | -1.8<br>$p=0.049$ |
| cg18942579<br>(chr17:57915773) | 4.0<br>[1.2; 8.6]% | 13.9<br>[6.3; 24.6]% | -9.9<br>$p=0.019$ | 2.6<br>[1.5; 8.8]% | 13.7<br>[6.0; 22.4]% | -11.1<br>$p=0.032$ |
| chr17:57915800 | *non-significant* | | | 8.3<br>[4.8; 17.1]% | 17.0<br>[9.0; 28.9]% | -8.7<br>$p=0.049$ |
| ***MIR21* gene body** | **Calcifications in the plaque shoulder** |  |  | **Fibrous cap rupture** |  |  |
|  | n=5 | n=10 |  | n=8 | n=7 |  |
| **Median** | 9.3<br>[8.8; 10.3]% | 6.8<br>[5.6; 8.8]% | 2.5<br>$p=0.044$ | 9.3<br>[7.6; 10.9]% | 6.3<br>[5.3; 8.3]% | 3.0<br>$p=0.044$ |
| cg02515217<br>(chr17:57918601) | 11.0<br>[9.0; 13.0]% | 6.2<br>[5.1; 7.4]% | 4.8<br>$p=0.044$ | 9.4<br>[5.7; 13.3]% | 6.7<br>[5.8; 7.8]% | 2.7<br>$p=0.044$ |

Note: $\beta$ , methylation level; $\Delta\beta$ , difference in methylation level between groups; n, sample size. $p$ , adjusted level of significance (FDR-corrected).

In contrast, methylation of cg02515217 and the median methylation level of the ***MIR21* gene body** were significantly higher in plaques with fibrous cap rupture or calcification in the plaque shoulder. We also observed significantly lower methylation of cg14399060 within the ***MIR10B*** region in plaques with thrombus on the plaque shoulder surface (Table 2). However, these findings require cautious interpretation because of the limited number of available samples (n ≤ 7) and the borderline statistical significance of the associations involving the *MIR21* gene body (*p*=0.044).

In addition, we identified moderate to strong correlations (|*rho*| = 0.4-0.9) between methylation levels at individual CpG sites in the ASPs and such quantitative histological indicators of atherosclerotic plaque instability as the number of inflammatory cells per field of view (Table 3).

**Table 3.** Moderate and strong correlations of methylation level of individual CpG sites in atherosclerotic plaque with individual <u>quantitative</u> histological indicators of plaque instability.

| Quantitative histological indicators of plaque instability | Region | CpG | Correlation coefficient and level of significance |
| --- | --- | --- | --- |
| Cap thickness, microm | **MIR127** | **chr14:101349509** | *rho* = -0.450; *p* = 0.016 |
|  |  | chr14:101349515 | *rho* = -0.510; *p* = 0.008 |
| Size of necrotic core, microm | **MIR127** | cg25063505 (chr14:101349286) | *rho* = -0.424; *p* = 0.035 |
|  |  | **chr14:101349509** | *rho* = -0.443; *p* = 0.018 |
|  |  | chr14:101349718 | *rho* = -0.520; *p* = 0.002 |
| Number of neutrophils (per field of view) | **MIR127** | chr14:101349561 | *rho* = -0.767; *p* = 0.010 |
|  |  | chr14:101349718 | *rho* = -0.673; *p* = 0.033 |
| Number of lymphocytes (per field of view) | **MIR127** | **chr14:101349643** | *rho* = -0.502; *p* = 0.006 |
|  |  | **chr14:101349784** | *rho* = 0.405; *p* = 0.018 |
| Number of macrophages (per field of view) | **MIR127** | **chr14:101349643** | *rho* = -0.571; *p* = 0.005 |
|  | **MIR127** | **chr14:101349784** | *rho* = 0.410; *p* = 0.037 |
| Number of foam cells (per field of view) | **MIR10B** | chr2:177015172 | *rho* = 0.538; *p* = 0.032 |
|  |  | chr2:177015208 | *rho* = 0.666; *p* = 0.005 |
|  | **MIR127** | chr14:101349856 | *rho* = 0.684; *p* = 0.042 |

Note: *p*, adjusted level of significance (FDR-corrected).
The inverse correlations are highlighted in gray. Strong correlations ( $|rho| > 0.7$ ) are shown in *italics*. CpG sites with consistent correlations across two plaque instability indicators are shown in **bold**.

Within the ***MIR127* region**, a strong correlation (|*rho*| = 0.7-0.9) was observed only between methylation of chr14:101349561 and the number of neutrophils per field of view (Table 3). No consistent methylation changes associated with any instability parameter were detected across the entire *MIR127* region or across consecutive CpG sites. Nevertheless, three CpG sites demonstrated concordant associations with two independent markers of plaque instability. Methylation of chr14:101349509 negatively correlated with both fibrous cap thickness and necrotic core size, whereas methylation of chr14:101349643 negatively correlated with number of macrophages and the number of lymphocytes. Conversely, methylation of chr14:101349784 showed positive correlations with both macrophage and lymphocyte number (Table 3).

We next compared DNA methylation levels in carotid ASP from patients with and without a history of acute cerebrovascular events (ACVE). Significant differences were detected for seven individual CpG sites within the ***MIR127*** region (*p* < 0.046) (Figure 6A, orange boxes), whereas the median methylation level of the entire ***MIR127*** region did not differ between the groups (*p* = 0.364) (Figure 6B). The observed methylation differences ranged from 2% to 4% and reached approximately 7% only for two adjacent CpG sites, chr14:101349509 and chr14:101349515 (Figure 6A, red box).

**Figure 6:**
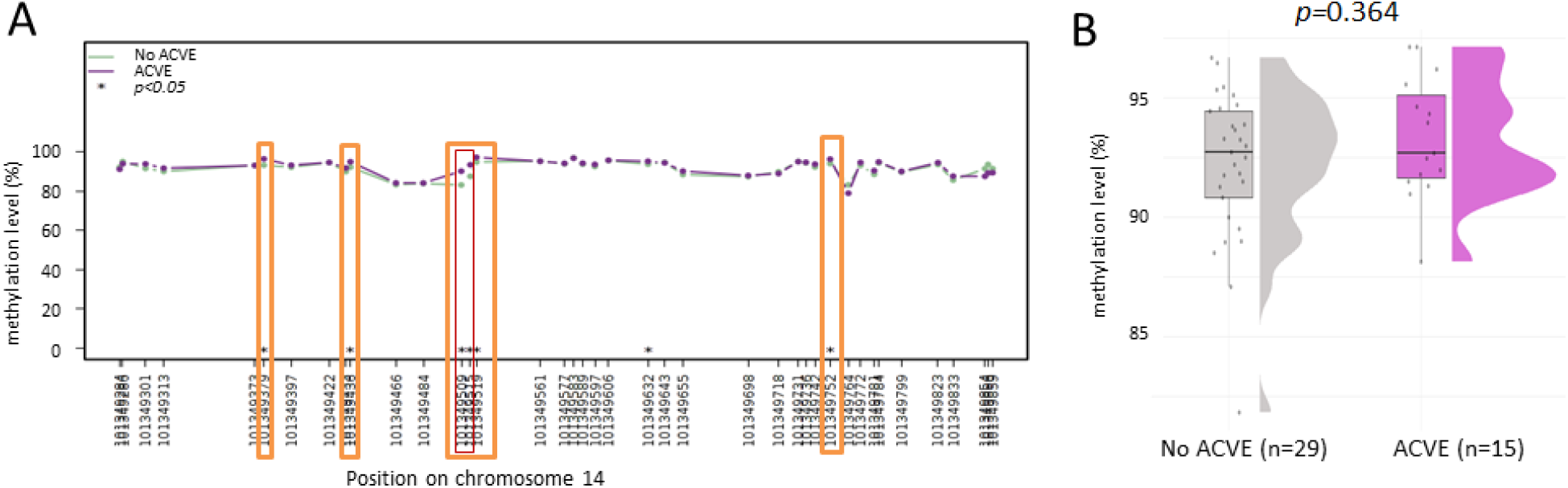
DNA methylation levels of *MIR127* gene in ASPs depending on the history of an acute cerebrovascular event (ACVE). (A) Methylation profile of CpG sites (The plot created in the online *Methylation plotter* tool [Mallona I. et al., 2014]. (B) Median DNA methylation levels for the entire *MIR127* gene region in ASP.

Plaques from patients with a history of ACVE exhibited significantly higher methylation levels at chr14:101349509 and chr14:101349515 (89.8 [85.0; 95.0]% and 94.8 [91.3; 96.5]%, respectively) than plaques from patients without previous ACVE (82.1 [77.0; 89.2]%, *p* = 0.001; and 87.9 [84.1; 91.7]%, *p* = 0.046, respectively). Notably, these same CpG sites also showed negative correlations with histological indicators of plaque instability. Specifically, methylation at both sites was inversely associated with fibrous cap thickness, whereas methylation at chr14:101349509 was additionally inversely associated with necrotic core size (Table 3).

At first glance, these findings appear contradictory, as increased methylation at chr14:101349509 is associated both with a history of ACVE and with reduced fibrous cap thickness (feature of plaque instability), while also correlating with a smaller necrotic core, whereas unstable plaques are generally characterized by large lipid core [Boswell-Patterson C.A. et al., 2023]. A possible explanation is that plaque vulnerability is commonly evaluated using the relative area occupied by the necrotic core (>25% of the plaque cross-sectional area) [Lovett J.K. et al., 2004; Gerhardt T. et al., 2022; Noonan J. et al., 2025], whereas in our study the necrotic core was measured as its absolute thickness (microm) on histological sections. Consequently, these two parameters may reflect different morphological aspects of plaque composition and are not necessarily directly comparable.

#### 2.4. Increased DNA methylation of a single CpG site within MIR100 E-box region in atherosclerotic plaque is associated with metabolic syndrome

The ***MIR100* E-box region**, a putative regulatory region of the *MIR100* gene, does not contain CpG sites annotated on Illumina methylation microarrays. Consequently, data regarding DNA methylation within this regulatory region remain limited.

Using BSAS, we assessed methylation at seven of the eight CpG sites within the ***MIR100* E-box region**, except for chr11:122025042 due to insufficient coverage. Among the analyzed sites, only chr11:122025143 demonstrated a significant association with clinical characteristics. Specifically, patients with metabolic syndrome exhibited higher methylation levels at this CpG site than patients without metabolic syndrome (92.2 [91.5; 92.6]% vs. 89.1 [86.7; 90.9]%, *p*=0.021) (Figure 7). No significant associations were identified between methylation at this or any other CpG site within the region and the remaining clinical characteristics examined.

**Figure 7:**
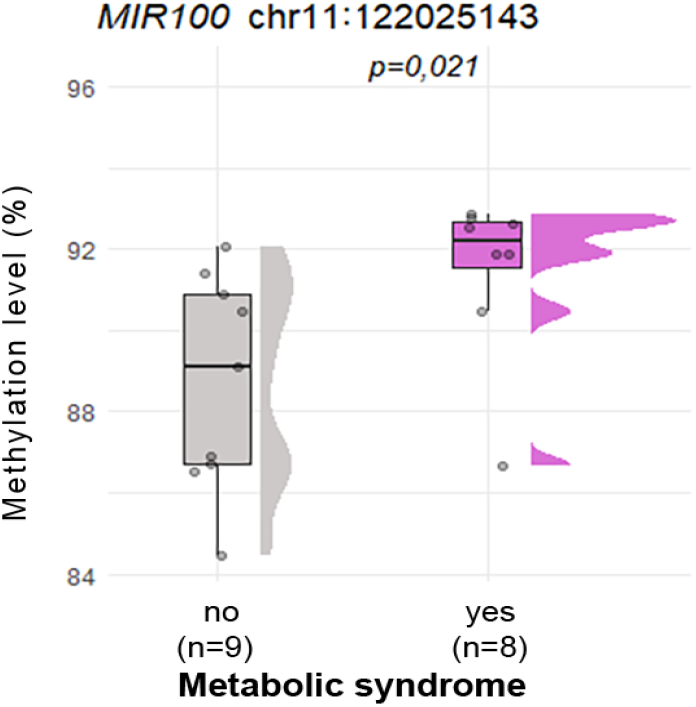
The methylation level of chr11:122025143 CpG site in the *MIR100* E-box region in the carotid artery ASPs depending on the presence of metabolic syndrome.

### 3. Methylation of microRNA genes in peripheral blood leukocytes in atherosclerosis

#### 3.1. Regions of MIR10B, MIR21 putative promoter, MIR21 CpG island and MIR100 E-box hypomethylated in blood of the patients with carotid atherosclerosis

The BSAS results revealed hypomethylation of individual CpG sites within the *MIR10B* region, *MIR21* CpG island and *MIR100* E-box region in PBP vs. PBC (Table 4). When examining the entire regions, a significant difference in median DNA methylation was observed only for the ***MIR21 putative promoter*** (Table 4, Figure 2).

**Table 4.** Statistically significant hypomethylation in CpG sites and entire *MIR21* putative promoter region in peripheral blood leukocytes of the patients with carotid atherosclerosis (PBP) compared with controls (PBC).

| Region | CpG site | $\beta$ in PBP | $\beta$ in PBC | $\Delta\beta$ |
| --- | --- | --- | --- | --- |
| *MIR10B* |  | (n=73) | (n=27) |  |
| | cg12215739 (chr2:177014966) | 10.2 [7.7; 16.4]% | 17.0 [10.7; 22.8]% | -6.8 ($p=0.016$) |
| | chr2:177015172 | 24.3 [19.0; 30.4]% | 27.7 [25.2; 31.5]% | -3.4 ($p=0.012$) |
| | chr2:177015208 | 19.1 [15.0; 27.9]% | 29.4 [17.1; 33.3]% | -10.3 ($p=0.047$) |
| *MIR21* putative promoter |  | (n=81) | (n=30) |  |
| | Median | 30.3 [21.8; 39.3]% | 36.8 [28.8; 46.0]% | -6.5 ($p=0.014$) |
| | chr17:57915172 | 0.7 [0.1; 1.8]% | 2.1 [0.6; 4.8]% | -1.4 ($p=0.005$) |
| | chr17:57915371 | 3.6 [1.8; 4.5]% | 6.9 [3.9; 11.2]% | -3.3 ($p=0.012$) |
| | cg12054453 (chr17:57915717) | 18.4 [10.5; 27.7]% | 25.9 [23.6; 32.4]% | -7.5 ($p=0.015$) |
| *MIR21* CpG island |  | (n=35) | (n=19) |  |
| | chr17:57916241 | 2.1 [1.2; 3.3]% | 4.2 [3.5; 6.8]% | -2.1 ($p=0.001$) |
| | chr17:57916306 | 3.6 [2.0; 7.3]% | 8.7 [6.7; 12.7]% | -5.1 ($p=0.001$) |
| | chr17:57916347 | 20.8 [16.8; 27.3]% | 29.6 [26.7; 33.2]% | -8.8 ($p<0.001$) |
| | chr17:57916385 | 49.1 [40.0; 56.1]% | 55.6 [52.0; 60.7]% | -6.5 ($p=0.025$) |
| | chr17:57916391 | 42.2 [30.7; 47.7]% | 45.8 [43.5; 49.8]% | -3.6 ($p=0.036$) |
| | chr17:57916416 | 63.3 [48.3; 68.2]% | 67.7 [64.5; 72.9]% | -4.4 ($p=0.038$) |
| | chr17:57916418 | 61.5 [47.6; 66.4]% | 66.7 [59.8; 72.5]% | -5.2 ($p=0.030$) |
| *MIR100* E-box |  | (n=18) | (n=17) |  |
| | chr11:122025072 | 95.9 [95.4; 97.2]% | 97.1 [96.6; 98.0]% | -1.2 ($p=0.041$) |

Note: $\beta$ , methylation level; $\Delta\beta$ , differences in methylation level between groups, $p$ , adjusted level of significance (FDR-corrected).

We next compared our BSAS data with the whole-blood EWAS dataset GSE69138 comprising 404 patients with ischemic stroke, distributed across 3 stroke subtypes: cardioembolic (n=127), lacunar (n=141), and stroke caused by large artery atherosclerosis (n=132) [Soriano-Tárraga C. et al., 2018].

Overall, the methylation patterns of the ***MIR21* putative promoter** region and ***MIR143*** regions were comparable between BSAS and EWAS (Figure 8). The largest discrepancies were observed for ***MIR127*** region, cg14399060 within ***MIR10B*** region and cg02515217 within the ***MIR21* gene body**. Compared with EWAS, BSAS showed lower methylation in the *MIR10B* region and *MIR21* gene body but higher methylation in *MIR127* region. These differences may partly reflect differences in disease stage, since samples in the current study were collected outside the acute cerebrovascular event (ACVE) period, whereas the external EWAS included patients sampled during the acute phase of stroke [Soriano-Tárraga C. et al., 2018].

**Figure 8:**
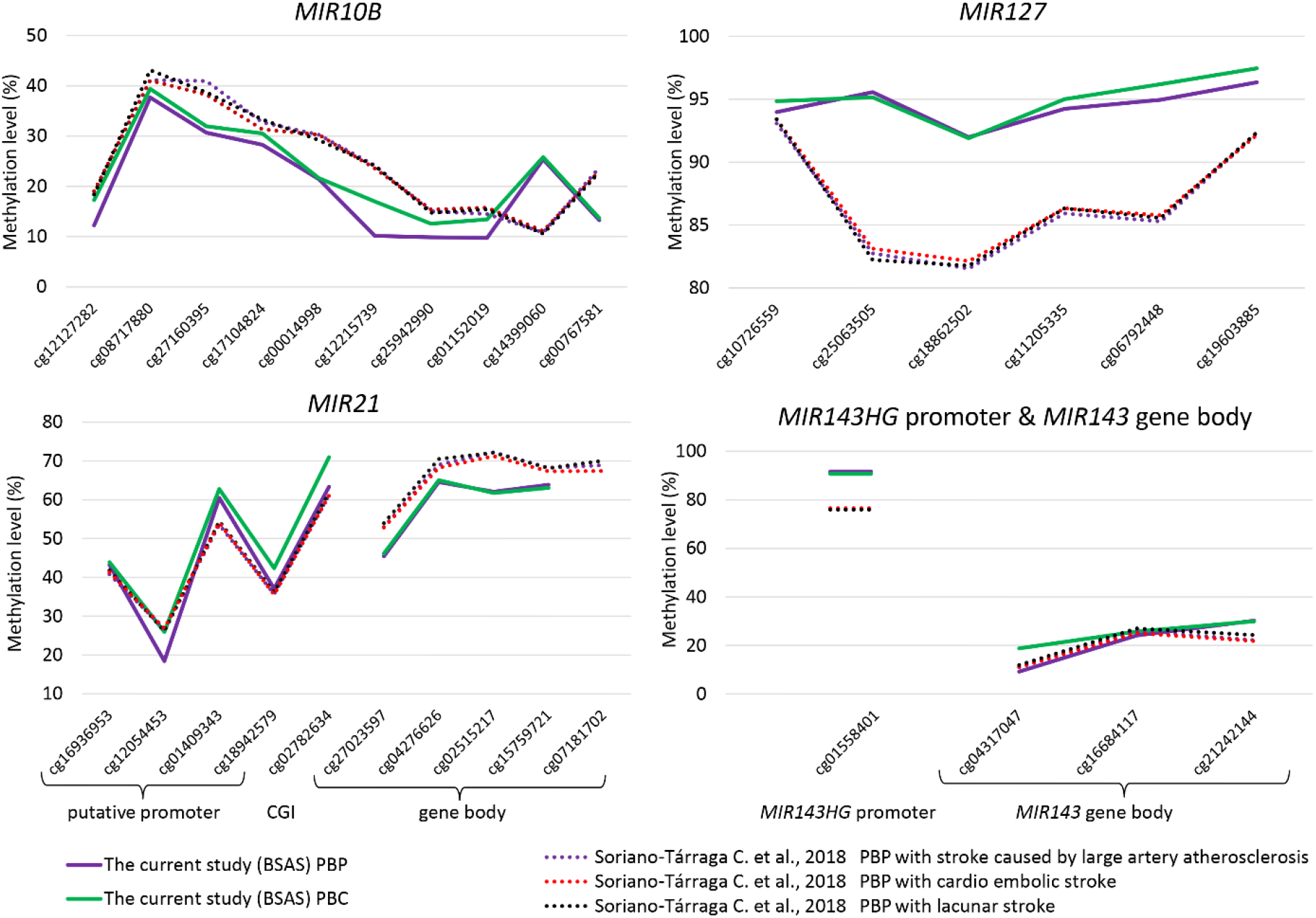
Comparison of DNA methylation at CpG sites annotated on Illumina methylation microarrays in the *MIR10B*, *MIR21*, *MIR127*, and *MIR143* regions in peripheral blood leukocytes in the current BSAS study and external EWAS dataset. BSAS, bisulfite amplicon sequencing; PBP, peripheral blood leukocytes of patients; PBC, peripheral blood leukocytes of controls

A history of ACVE had little influence on leukocyte DNA methylation in our BSAS results. Significant differences were detected only for two CpG sites, both with small effect sizes (2.0-3.1%): cg01152019 (chr2:177015044) within *MIR10B* region showed lower methylation (8.1 [6.8; 11.0]% vs 11.2 [8.3;13.8]%, *p*=0.044), whereas chr14:101349436 within *MIR127* region showed higher methylation in patients with previous ACVE (96.4 [95.0; 97.4]% vs 94.4 [92.2; 96.5]%, *p*=0.033). No significant associations were observed for the remaining analyzed regions, although several comparisons were limited by the small number of patients with previous ACVE.

#### 3.2. DNA methylation level of the MIR10B and MIR143 genes in blood leukocytes of patients with atherosclerosis is associated with blood lipids and degree of carotid stenosis

Next, we analyzed the associations between the DNA methylation level in the studied regions of microRNA in the in leukocytes of patients with atherosclerosis and pathogenetically relevant traits of atherosclerosis.

The analysis revealed strong negative correlations (rho <-0.7) for chr2:177015208 in the ***MIR10B*** gene with serum glucose, total cholesterol and atherogenic index (Table 5). For cg12215739 (chr2:177014966) we observed strong negative correlation with atherogenic index and moderate negative correlations (rho =-0.4…-0.69) with total cholesterol level. Both cg12215739 (chr2:177014966) and chr2:177015208 showed moderate negative correlations with low-density lipoprotein levels and with the degree of carotid artery stenosis. There were also single correlations of methylation of other CpG sites in ***MIR10B* gene region** with stenosis degree or total cholesterol levels (Table 5).

**Table 5.**
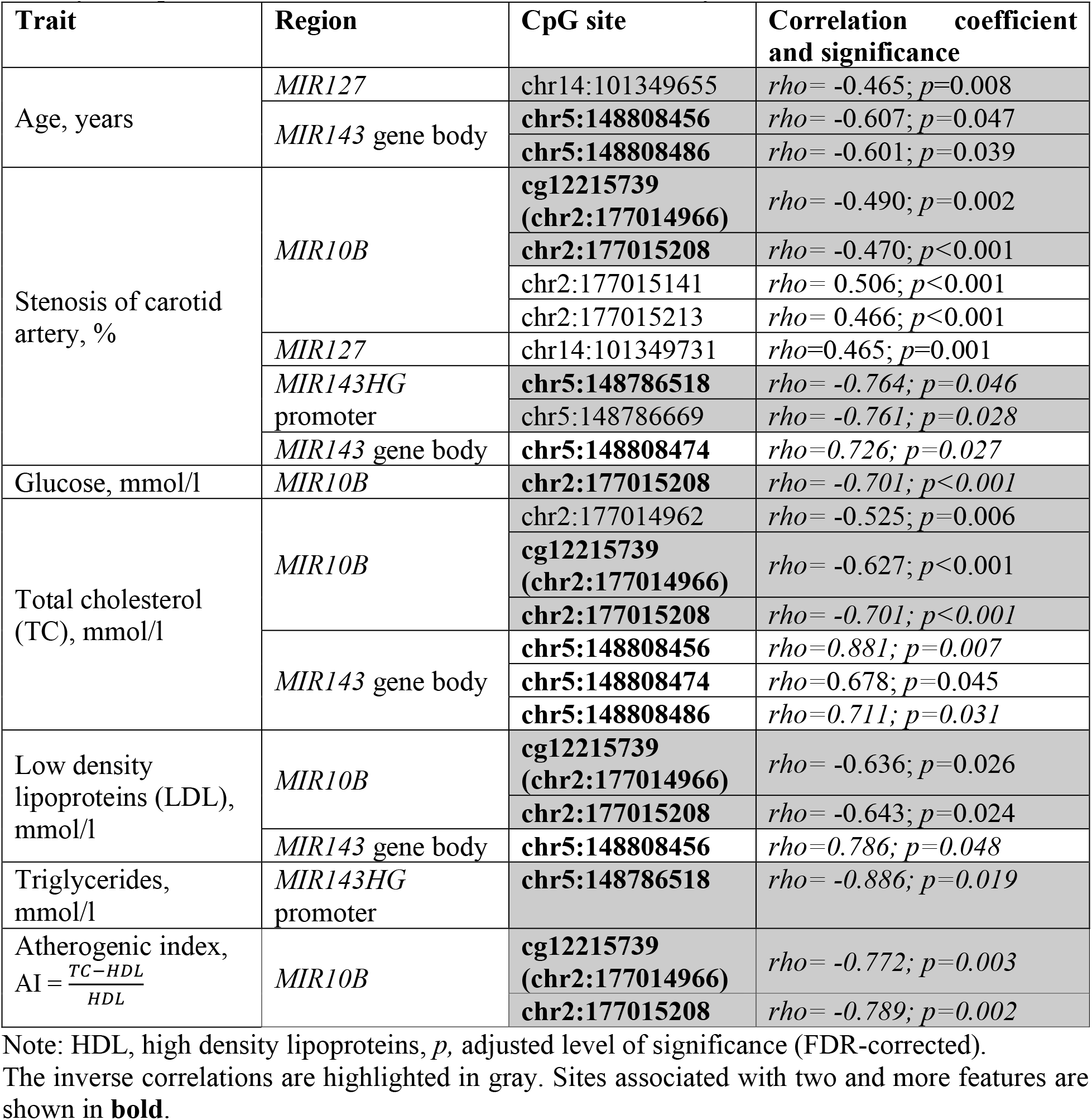
Correlations between DNA methylation of individual CpG sites in peripheral blood leukocytes of patients with carotid atherosclerosis and clinically relevant traits.

The strong negative correlations with degree of carotid artery stenosis and triglycerides levels were revealed for methylation level of 2 CpG sites in ***MIR143HG* promoter**, and the chr5:148786518 showed strong correlation with both traits. At the same time, the CpG sites of ***MIR143* gene body** showed positive correlations with stenosis degree and parameters of lipid metabolism (total cholesterol and low-density lipoproteins). The moderate negative correlations with patients age were also noted for the CpG sites of ***MIR143* gene body** (Table 5).

None of CpG sites in the ***MIR127* gene** region showed significant correlation with more than one pathogenetically relevant trait (Table 5). Also, none of the correlations of DNA methylation for leukocytes of patients with atherosclerosis were confirmed for the leukocytes of control group (*p* > 0.084).

#### 3.3. MIR21 gene body and MIR143 gene body methylation levels are associated with cellular composition of blood

Peripheral blood is a frequently used material for analysis, but leukocyte composition may influence whole-blood DNA methylation. So, we estimated cell-type proportions in an external EWAS dataset (GSE246337) and evaluated correlations between methylation of the analyzed CpG sites and blood cell composition. One DNAm reference matrix designed for adult whole blood in EpiDISH algorithm can estimate fractions for 7 immune cell types (B-cells, NK-cells, CD4T and CD8T-cells, Monocytes, Neutrophils and Eosinophils) [Teschendorff A.E. et al. 2017]. The blood cell estimation of external EWAS (GSE246337) was evaluated by 28 CpG sites of microRNA genes annotated on Illumina methylation microarrays (Supplementary Table 14).

CpG sites (cg16684117 and cg21242144) in the ***MIR143* gene body** showed strong positive correlations with the percentage of B-cells and CD4T-cells (Figure 9). CpG sites within ***MIR21* and *MIR127* genes** were positively associated with the percentage of neutrophils, with the association being particularly pronounced for the *MIR21* locus (Figure 9). DNA methylation within the ***MIR10B*** region exhibited only weak or negligible correlation with main cell types in peripheral blood (Figure 9).

**Figure 9:**
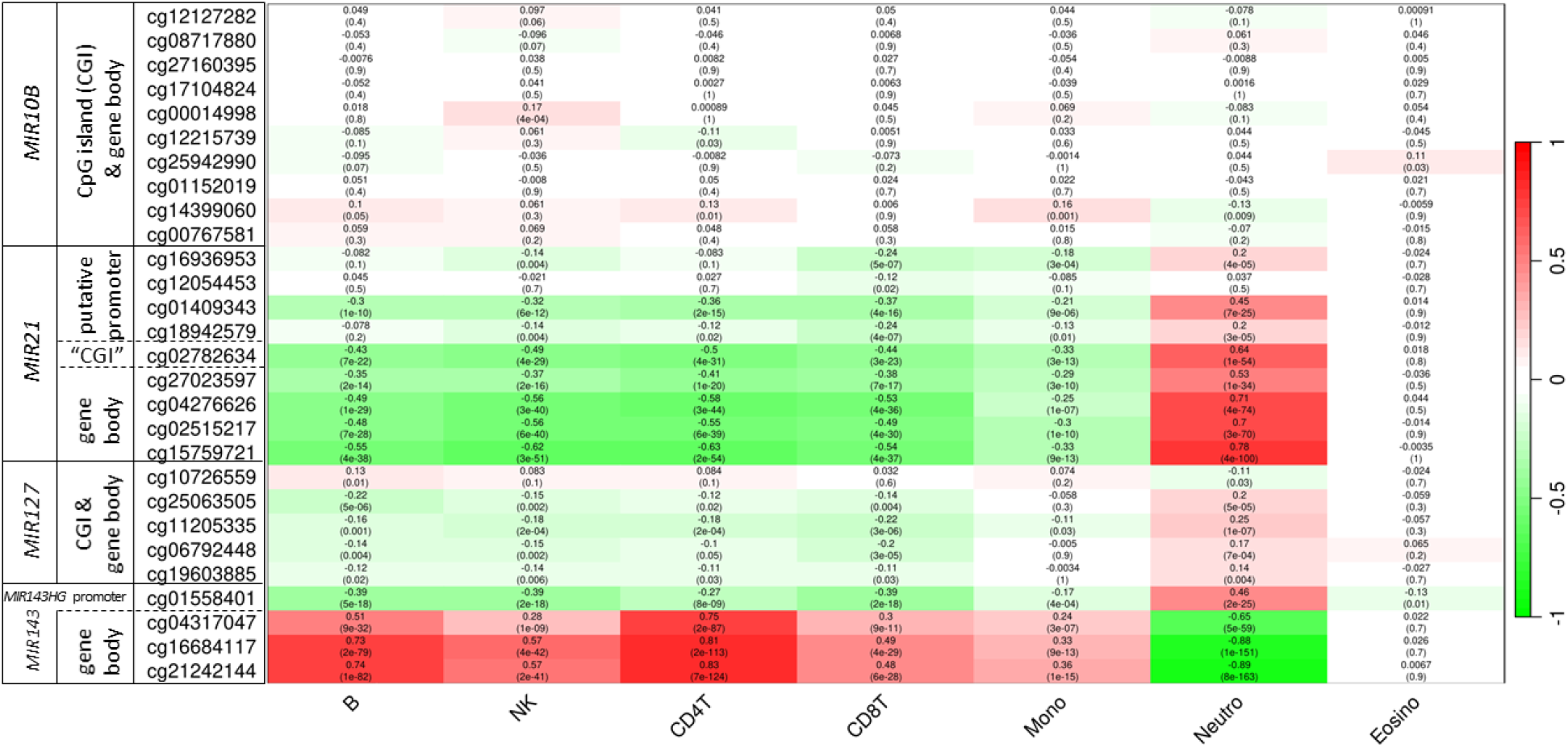
Heatmap of correlations between DNA methylation of analyzed CpG sites and the main blood cell types in peripheral blood – B-cells, NK, CD4T-cells, CD8T-cells, monocytes (Mono), neutrophils (Neutro), eosinophils (Eosino).

Interestingly, in the case of vascular wall, these sites showed a strong positive correlation with the percentage of SMCs (Figure 4). The methylation status of the ***MIR143HG*** promoter site cg01558401 correlated significantly with peripheral blood neutrophils and IC infiltration in carotid plaques (Figures 4, 9), indicating that its variation is driven by cellular heterogeneity.

#### 3.4. MIR21 CpG island methylation in peripheral blood strongly reflects its methylation status in carotid atherosclerotic plaques

Using our BSAS results, we compared the methylation of the studied microRNA regions in atherosclerotic plaques of patients and in their peripheral blood (ASP vs PBP). The significant positive correlations observed for the median DNA methylation level of ***MIR10B* region** (*rho* = 0.336, *p* = 0.009), ***MIR21* putative promoter** (*rho* = 0.432, *p* < 0.001), ***MIR21* CpG island** (*rho* = 0.722, *p* < 0.001) and ***MIR143* gene body** (*rho* = 0.482, *p* = 0.038) (Supplementary Table 01).

The strongest correlations (*rho* > 0.7) were observed for cg14399060 within ***MIR10B* gene**, (chr2:177015070, *rho* = 0.841, *p* < 0.001) and four consecutive CpG sites within the ***MIR21* CpG island**: chr17:57916598 (*rho* = 0.745, *p* < 0.001), chr17:57916641 (*rho* = 0.745, *p* < 0.001), cg02782634 (chr17:57916643, *rho* = 0.702, *p* < 0.001) and chr17:57916647 (*rho* = 0.731, *p* < 0.001). Notably, cg14399060 was also associated with thrombus on the plaque surface (Table 2) and differed in peripheral blood leukocytes between our BSAS data and the external stroke EWAS dataset [Soriano-Tárraga C. et al., 2018].

These findings suggest that peripheral blood leukocytes, as an easily accessible source for DNA analysis, may provide a surrogate material for indirect assessing DNA methylation in carotid plaques. Four CpG sites and the entire ***MIR21* CpG island** region seem to be the most suitable for this purpose. However, because these analyses were based on only 13 paired ASP-PBP samples, validation in larger cohorts with assessment of diagnostic performance is required.

## Discussion

DNA methylation of microRNA genes has been investigated predominantly in cancer, whereas studies addressing vascular tissues and peripheral blood leukocytes in atherosclerosis remain scarce. Consequently, the epigenetic regulation of microRNA genes in atherosclerosis is still poorly understood. To our knowledge, this study represents one of the most comprehensive targeted analyses of DNA methylation across multiple microRNA gene regions in advanced carotid atherosclerosis. A major strength of the present study is the simultaneous analysis of vascular tissues and peripheral blood leukocytes obtained from the same patients with carotid atherosclerosis. In addition to microRNA gene bodies, we investigated the regulatory elements, including CpG islands, promoter regions, and a transcription factor binding site.

Our results demonstrate that advanced carotid atherosclerosis is characterized by a moderate DNA hypomethylation within the vascular wall, specifically affecting the *MIR10B* and *MIR127* regulatory regions, the *MIR100* E-box element, and the gene bodies of *MIR21* and *MIR143*, which parallelly mirrors the hypomethylation of individual CpG sites within the *MIR10B* region, *MIR21* putative promoter, *MIR21* CpG island, and *MIR100* E-box region observed in peripheral blood with a moderate effect size (12% and less).

However, adjustment for estimated cell composition substantially altered these associations. After accounting for SMCs and ICs in vascular tissue, methylation changes in the *MIR100* E-box region and the *MIR127* remained independently associated with carotid atherosclerosis, suggesting that these loci primarily reflect disease-related molecular alterations rather than differences in cellular composition.

The deconvolution analysis of vascular tissue revealed a striking mirror-image dichotomy, where higher DNA methylation within the *MIR10B* and *MIR127* (except cg25063505) loci and the *MIR143* gene body serves as an epigenetic signature of SMCs, whereas methylation of the *MIR21* and *MIR143HG* promoters reflect IC infiltration. The opposite correlation of the DNA methylation of the *MIR143HG* promoter/*MIR143* gene body with ICs/SMCs likely reflects the cell-specific epigenetic regulation of gene. It was suggested that DNA methylation of microRNA gene body may foster the transcription of microRNA [Huan T. et al., 2020].

Peripheral blood DNA methylation profiles of the most analyzed microRNA genes are profoundly shaped by leukocyte composition. While the methylation variations within the *MIR21, MIR127* and *MIR143* loci are strongly confounded by specific IC fractions, mainly neutrophils, the *MIR10B* region remains robust and unaffected by cellular shifts. These findings emphasize the importance of accounting for cellular heterogeneity in epigenetic studies of both vascular and blood tissues.

Among the investigated genes, *MIR127* has been the most extensively studied in atherosclerosis. Previous analyses of femoral atherosclerotic plaques demonstrated hypomethylation of the 14q32 locus containing *MIR127* [Aavik E. et al., 2015]. However, only relative fold changes rather than absolute methylation levels were reported, limiting direct comparison with our results.

Another study also described relatively low (less than 60%) methylation across several regions within the 14q32 locus in different vascular tissues: lower limb arteries and veins, upper limb veins and mammary arteries [Goossens E.A.C. et al., 2019]. Unfortunately, the study was not conducted on the carotid arteries, and regions of the 14q32 locus also differed in location from the region analyzed in our study: one of the nearest regions was located 57 Kb upstream and other 22 Kb downstream *MIR127* gene body and therefore they did not overlap the CpG island examined in our study.

In contrast, we observed consistently high methylation (>80%) across nearly all CpG sites within the *MIR127* region in all types of studied tissues, including affected and intact carotid arteries. Notably, publicly available microarray datasets (GSE66500, GSE149759, GSE69138) demonstrated similarly high methylation levels for Illumina-annotated CpG sites within this region (Figures 3 and 8), supporting the conclusion that high constitutive methylation is a characteristic feature of the *MIR127* region despite minor differences between studies and analytical platforms.

Published data on DNA methylation of the *MIR100* regulatory region are extremely limited. Chen D. et al. (2014) reported hypermethylation of the *MIR100HG* (miR-100 host gene) in breast cancer [Chen D. et al., 2014]. However, the genomic coordinates of the affected region were not specified, preventing direct comparison with our data. We identified a low-magnitude increase in methylation at a single CpG site (chr11:122025143, GRCh37/hg19) within the *MIR100* E-box region in plaques from patients with metabolic syndrome.

Because the ZEB1 transcription factor binds to this E-box and enhances miR-100 expression [Chen D. et al., 2014], increased methylation of even single CpG at this regulatory element may interfere with transcription factor binding and reduce miR-100 expression. This interpretation is consistent with previous reports demonstrating reduced miR-100 expression in macrophage-rich and lipid-rich atherosclerotic plaques compared with lesions with a low macrophage or lipid content [Pankratz F. et al., 2018]. Also circulating miR-100 was significantly lower in obese normoglycemic subjects and subjects with type 2 diabetes compared to healthy individuals, and in visceral adipose tissue, expression of miR-100 was lower in obese subjects with diabetes compared to obese subjects without diabetes [Pek S.L. et al., 2016]. Furthermore, experimental studies suggest a protective role of miR-100 against high-fat diet-induced metabolic syndrome [Smolka C. et al., 2021], supporting the potential biological relevance of the methylation changes identified in the present study.

For *MIR10B* region, we observed significant decrease in the DNA methylation level in the carotid arteries affected by atherosclerosis. This is consistent with the results of our previous research, where methylation levels of four CpG-sites located within the *MIR10B* gene body and CpG island were decreased in coronary atherosclerotic plaques in comparison with the other vascular tissues [Nazarenko M.S. et al., 2015]. These observations indicate that reduced methylation of the *MIR10B* locus may represent a common epigenetic feature of advanced atherosclerosis across different vascular beds.

Although *MIR21* methylation has been extensively investigated in cancer and autoimmune diseases, information regarding its methylation (both in the putative promoter region and the gene body) in vascular tissues or peripheral blood leukocytes from patients with atherosclerosis has been lacking. Likewise, data on methylation of the *MIR143HG* promoter and the *MIR143* gene body remain limited. There was one study that analyzed the DNA methylation level of *MIR145* gene and *MIR143HG* promoter by pyrosequencing in SMCs of saphenous veins from patients with type 2 diabetes mellitus, but it was unable to obtain reliable measurements for *MIR143HG* promoter [Riches K. et al., 2017]. Conversely, technical limitations prevented assessment of *MIR145* methylation in the present study. Previous genome-wide analyses have identified several hypermethylated regions within the long non-coding RNA *CARMN* (*MIR143HG*) gene, which overlaps the sequences encoding miR-145 and miR-143, although the exact genomic coordinates of these regions were not reported, precluding direct comparison [Ehrlich K.C. et al., 2019].

Beyond identifying differential methylation between vascular tissues, we demonstrated significant associations between DNA methylation and clinically relevant characteristics of plaque instability. Hypomethylation of individual CpG sites within *MIR10B* and the *MIR21* putative promoter was associated with the presence of thrombi on the plaque surface, intraplaque hemorrhage, and the presence of foam cells. In addition, two CpG sites within the *MIR127* region (chr14:101349509 and chr14:101349515, GRCh37/hg19) exhibited increased methylation in patients with a history of ACVE and were also associated with histological indicators of plaque instability.

Only one previous study has investigated methylation of microRNA genes in peripheral blood leukocytes from patients with cardiovascular disease. That genome-wide study on Human Methylation450 BeadChip microarray identified a nominal association for cg16936953 (chr17:57915665) within the *MIR21* putative promoter in myocardial infarction (*p*=0.049); however, significance was lost after correction for multiple testing [Ek W.E. et al., 2016]. In contrast, this CpG site was not differentially methylated between leukocytes of patients with atherosclerosis and controls in our cohort, suggesting that methylation changes in *MIR21* may differ between carotid atherosclerosis and myocardial infarction or may depend on disease stage.

Overall, our BSAS and lab-internal EWAS data showed good agreement with external EWAS datasets regarding both methylation levels and methylation patterns of *MIR10B*, *MIR21*, *MIR127*, and *MIR143* for leukocytes [Soriano-Tárraga C. et al., 2018] and for vascular tissues [Zaina S. et al., 2014; Zaina S. et al., 2015; Li J. et al., 2021]. Some discrepancies between studies likely reflect methodological differences between bisulfite amplicon sequencing and microarray-based approaches, as well as biological differences among vascular beds and patient populations. Nevertheless, the overall concordance between datasets supports the robustness of the identified methylation patterns.

In our study the methylation level of some CpG sites within *MIR10B*, *MIR143HG* promoter, and *MIR143* gene body in leukocytes of patients with atherosclerosis exhibited significant associations with blood lipids parameters and degree of carotid stenosis. Furthermore, the correlation between methylation of *MIR10B*, *MIR21*, and *MIR127* in ASP and histological indicators of plaque instability and acute cerebrovascular events support their clinical relevance in predicting plaque rupture.

As thoroughly reviewed by Breton et al. (2017), small-magnitude effect sizes (1-5% differences) in DNA methylation are highly characteristic of complex, non-malignant chronic diseases analyzed in peripheral blood. Such low-level shifts are functional and robustly validate gene-expression changes [Breton C.V. et al., 2017]. Thus, we believe that the level of DNA methylation of the *MIR10B*, *MIR21*, *MIR127*, and *MIR143* gene regions in the PBP may serve as biomarker for the development of carotid artery atherosclerosis. Importantly, we also demonstrated strong correlations between DNA methylation levels in peripheral blood leukocytes and matched atherosclerotic plaques, particularly for the *MIR21* CpG island. These findings suggest that selected CpG sites within this region may serve as surrogate markers of plaque methylation and provide a basis for developing minimally invasive epigenetic biomarkers. Nevertheless, this observation requires validation in larger prospective cohorts with formal evaluation of diagnostic performance, including sensitivity, specificity, and receiver operating characteristic analyses.

The study has several limitations that should be considered when interpreting the results. First, the relatively modest sample size reduced statistical power, particularly for subgroup analyses involving acute cerebrovascular events or specific histological features. Second, only a limited number of microRNA genes and regulatory regions were investigated, while technical limitations prevented analysis of the *MIR100* gene body and *MIR145* gene body. Finally, although we accounted for vascular cellular heterogeneity, functional experiments linking DNA methylation changes with microRNA expression and downstream biological effects were beyond the scope of this study. So, broader epigenome-wide studies might reveal additional methylation changes relevant to carotid atherosclerosis.

Despite these limitations, it is worth noting that in this study, we analyzed the methylation level of both microRNA genes themselves and their regulatory elements, and the localization of these regulatory elements was obtained through a search of open sources. This methylation analysis took into account histological parameters of plaque instability, cellular heterogeneity, and the use of EWAS data. For the first time, it has been shown that the regulatory elements of microRNA genes in advanced atherosclerosis are predominantly hypomethylated, in contrast to cancer. Moreover, correction for cellular heterogeneity identified that *MIR100* and *MIR127* methylation changes most likely reflecting disease-associated molecular alterations rather than shifts in vascular cell composition. Finally, the strong correspondence between methylation profiles in plaques and peripheral blood supports the potential use of four CpG sites within the *MIR21* CpG island, along with the entire *MIR21* CpG island region as minimally invasive surrogate biomarkers of plaque epigenetic status.

## Conclusion

This study demonstrates tissue-specific moderate DNA methylation alterations featuring hypomethylation in microRNA regulatory regions associated with advanced carotid atherosclerosis.

DNA methylation profiles of the analyzed regions of microRNA genes are profoundly shaped by cellular composition in both vascular tissues and blood. After adjustment for vascular cellular heterogeneity, methylation changes within the *MIR100* and the *MIR127* genes remained independently associated with carotid atherosclerosis. DNA methylation within the *MIR10B* exhibited weak or negligible correlation with blood cell composition.

In carotid atherosclerotic plaques methylation changes within *MIR100* were associated with metabolic syndrome, while alterations in *MIR10B, MIR21,* and *MIR127* were linked to histological plaque instability and history of acute cerebrovascular events. In blood, *MIR10B* and *MIR143* methylation were associated with lipid metabolism and carotid stenosis, whereas methylation of the *MIR21* CpG island showed strong correspondence between blood and plaque, supporting its potential as a surrogate biomarker in future studies.

## Data Availability

All data produced in the present work are contained in the manuscript and supplementary material

https://www.ncbi.nlm.nih.gov/geo/query/acc.cgi?acc=GSE46394

https://www.ncbi.nlm.nih.gov/geo/query/acc.cgi?acc=GSE149759

https://www.ncbi.nlm.nih.gov/geo/query/acc.cgi?acc=GSE69138

https://www.ncbi.nlm.nih.gov/geo/query/acc.cgi?acc=GSE246337

https://www.ncbi.nlm.nih.gov/geo/query/acc.cgi?acc=GSE66500

## Acknowledgments

During manuscript preparation, the authors used OpenAI ChatGPT (GPT-5.6 Pro; OpenAI) for English-language and stylistic editing. The authors reviewed and verified all edits and take full responsibility for the final content.

