## Supplementary Methods for "DNA methylation alterations in *MIR10B*, *MIR21*, *MIR100*, *MIR127* and *MIR143* genes associated with advanced carotid artery atherosclerosis: a targeted bisulfite sequencing study of vascular tissues and peripheral blood"

### *Supplementary Material*

### Supplementary Methods

**1. Study participants and control group.**

**Table SM1.** Clinical characteristics of the study groups included

**2. Histological examination of carotid atherosclerotic plaques**

**Table SM2.** Morphological characteristics of carotid atherosclerotic plaques (N=16) included in the Infinium MethylationEPIC BeadChip (Illumina) microarray study

**Table SM3.** Clinical characteristics of the groups included in the Infinium MethylationEPIC BeadChip (Illumina) microarray study

**3. Selection of miRNA and *MIR* gene regions for DNA methylation analysis.**

**Table SM4**. Expression of microRNAs associated with carotid atherosclerotic plaque instability in humans

**Table SM5.** Candidate genes and regions for the analysis of miRNA regulatory elements

**Table SM6**: Studied regions and primer structure

**Supplementary Figure 1. Determining the location of regulatory elements of the *MIR21* gene.**

**Supplementary Figure 2. Determining the location of regulatory elements of the *MIR143* and *MIR145* genes.**

**4. DNA methylation analysis by Bisulfite Amplicon Sequencing (BSAS)**

**4.1 Primer design for BSAS**

**4.2. Amplification of target DNA regions**

**4.3 Equimolar pooling and purification of amplicons**

**4.4. DNA library preparation and sequencing**

**4.5. Bioinformatic processing of the results**

**References**

#### 1 Study participants and control group.

This study included 92 patients with advanced carotid atherosclerosis and 32 control participants without symptoms of cerebrovascular disease (**Table SM1**). Participants were recruited at the Research Institute of Complex Problems of Cardiovascular Diseases (Kemerovo, Russia) and the Cardiology Research Institute (Tomsk National Research Medical Center, Tomsk, Russia) in 2014-2017. All participants provided written informed consent in accordance with the principles of the Declaration of Helsinki.

Exclusion criteria were non-Caucasian ethnicity or first-degree relationship with another study participant, age less than 40 years, presence of acute severe comorbid somatic pathology or exacerbation of a chronic disease; or refusal to participate.

**Table SM1.** Clinical characteristics of the study groups included

| Parameter | Patients with clinically advanced carotid atherosclerosis (n=92) | Control group of healthy individuals (n=32) | *p* value |
|---|---|---|---|
| **Clinical parameters** | | | |
| Men: Women | 64:28 | 19:13 | 0.402 |
| Age, years (M [Q1; Q3]) | 64.0 [60.0; 69.0] | 67.0 [58.8; 71.8] | 0.196 |
| Body mass index, kg/m$^2$ (M [Q1; Q3]) | 28.4 [25.9; 31.6] | 26.2 [23.4; 29.1] | **0.012** |
| Metabolic syndrome [n (%)] | 61 (66.3%) | *no data* | — |
| Waist circumference, cm (M [Q1;Q3]) | 101.0 [93.0; 113.0] | *no data* | — |
| History of angina [n (%)] | 61 (66.3%) | 0 (0.0%) | **0.002** |
| Angina functional class [n (%)]<br>- I FC<br>- II FC<br>- III FC | <br>12 (13.0%)<br>35 (38.0%)<br>14 (15.2%) | *no data* | — |
| History of myocardial infarction [n (%)] | 36 (39.1%) | 2 (6.3%) | **<0.001** |
| History of coronary artery disease [n (%)] | 75 (81.5%) | 5 (15.6%) | **<0.001** |
| Duration of coronary artery disease, years (M [Q1;Q3]) | 2.5 [1.0; 9.8] | *no data* | — |
| History of chronic heart failure [n (%)] | 76 (82.6%) | 1 (3.1%) | **<0.001** |
| Chronic heart failure functional class [n (%)]<br>- I FC<br>- II FC<br>- III FC | <br>6 (6.5%)<br>63 (68.5%)<br>8 (8.7%) | *no data* | — |
| History of TIA or stroke [n (%)] | 26 (28.3%) | 0 (0.0%) | **<0.001** |
| Hypertension [n (%)] | 90 (97.8%) | 15 (46.9%) | **<0.001** |
| Duration of hypertension, years (M [Q1; Q3]) | 13.5 [7.3; 20.0] | *no data* | — |
| Hypercholesterolemia [n (%)] | 63 (68.5%) | 5 (15.6%) | — |
| Smoking [n (%)]<br>including former smokers | 60 (65.2%)<br>13 (14.1%) | 11 (34.4%)<br>0 (0.0%) | **0.002**<br>0.020 |
| Diabetes mellitus [n (%)] | 22 (23.9%) | 3 (9.4%) | 0.123 |
| Chronic limb ischemia [n (%)] | 19 (20.7%) | *no data* | — |
| **Vascular ultrasound parameters (M [Q1; Q3])** | | | |
| Carotid artery stenosis, % | 85 [80; 95] | 28 [24; 45] | **<0.001** |
| Intima-media complex thickness, mm | *no data* | 0.8 [0.7; 0.9] | — |

**Table SM1 (continued)**

| Parameter | Patients with clinically advanced carotid atherosclerosis (n=92) | Control group of healthy individuals (n=32) | *p* value |
|---|---|---|---|
| **Laboratory data (M [Q1; Q3])** | | | |
| Glucose, mmol/l | 5.8 [5.3; 7.3] | 5.4 [5.1; 5.7] | **0.017** |
| Total cholesterol, mmol/L | 5.3 [4.0; 6.3] | 5.2 [4.7; 5.7] | 0.898 |
| Triglycerides, mmol/L | 1.5 [1.3; 2.0] | 1.3 [1.0; 1.5] | 0.077 |
| LDL, mmol/L | 2.9 [2.0; 3.7] | 3.1 [2.9; 3.7] | 0.641 |
| HDL, mmol/L | 1.0 [1.0; 1.1] | 1.4 [1.1; 1.6] | **0.017** |
| Atherogenic index (AI = (Total cholesterol - HDL)/HDL) | 4.8 [2.4; 5.3] | 2.7 [2.3; 3.3] | 0.140 |
| **Medications [n (%)]** | | | |
| Aspirin | 65 (70.7%) | 0 (0.0%) | **<0.001** |
| Clopidogrel | 25 (27.2%) | 1 (3.1%) | **<0.001** |
| Warfarin | 10 (10.9%) | 0 (0.0%) | 0.059 |
| Beta-blockers | 57 (62.0%) | 5 (15.6%) | **<0.001** |
| Angiotensin converting enzyme inhibitors | 44 (47.8%) | 2 (6.3%) | **<0.001** |
| Angiotensin II receptor blockers | 27 (29.3%) | 5 (15.6%) | 0.160 |
| Calcium channel blockers (dihydropyridine) | 47 (51.1%) | 1 (3.1%) | **<0.001** |
| Calcium channel blockers (non-dihydropyridine) | 9 (9.8%) | 0 (0.0%) | 0.110 |
| Digoxin | 11 (12.0%) | 0 (0.0%) | **0.035** |
| Loop diuretics | 14 (15.2%) | 0 (0.0%) | **0.020** |
| Thiazide diuretics | 18 (19.6%) | 2 (6.3%) | 0.096 |
| Spironolactone | 14 (15.2%) | 0 (0.0%) | **0.020** |
| Nitrates | 9 (9.8%) | 0 (0.0%) | 0.110 |
| Statins | 67 (72.8%) | 1 (3.1%) | **<0.001** |
| Cordarone | 17 (18.5%) | 0 (0.0%) | **0.006** |
| Insulin | 9 (9.8%) | 0 (0.0%) | 0.110 |
| Metformin | 16 (17.4%) | 1 (3.1%) | 0.039 |

**Abbreviations:** n, number of individuals in the group; M, median; Q1, first quartile (25th percentile); Q3, third quartile (75th percentile).
FC, functional class; HDL, high density lipoproteins; LDL, low density lipoproteins; TIA, transient ischemic attack.

#### 2 Histological examination of carotid atherosclerotic plaques

Histological examination was performed for 56 carotid ASP samples (60.7 %). Tissue fragments were fixed in 10% neutral formalin, processed in a Shandon Excelsior ES Tissue Processor (Thermo Scientific, USA), and embedded in paraffin. Serial 5-µm paraffin sections (5-6 microns) were prepared using a Microm HM 430 microtome (Thermo Scientific, USA), stained with hematoxylin-eosin and Van Gieson's stain, examined using an Axio Lab.A1 microscope (Carl Zeiss, Germany), and imaged with a MIRAX MIDI slide scanner (Carl Zeiss, Germany). Morphometric analysis was performed using ImageJ (NIH, USA). Plaques were classified according to the American Heart Association system (AHA) [Stary et al. 1995].

**Table SM2.** Morphological characteristics of carotid atherosclerotic plaques (N=16) included in the Infinium MethylationEPIC BeadChip (Illumina) microarray study

| | |
|---|---|
| **Assessment of the atherosclerotic plaque cap** | |
| Cap thickness, microm (M [Q1;Q3]) | 308.5 [236.2; 468.6] |
| Thinnings [n (%)] | 11 (68.8%) |
| Ruptures [n (%)] | 8 (50.0%) |
| Erosions [n (%)] | 6 (37.5%) |
| Ulcerations [n (%)] | 5 (31.3%) |
| Calcifications [n (%)] | 1 (6.3%) |
| **Assessment of the atherosclerotic plaque core** | |
| Core size, microm (M [Q1;Q3]) | 1803.5 [1087.9; 2234.8] |
| Calcifications [n (%)] | 4 (25.0%) |
| **Assessment of the atherosclerotic plaque shoulder** | |
| Ruptures [n (%)] | 7 (43.8%) |
| Thrombus [n (%)] | 3 (18.8%) |
| Hemorrhages [n (%)] | 7 (43.8%) |
| Calcifications [n (%)] | 2 (12.5%) |
| **Assessment of inflammatory infiltration in atherosclerotic plaque** | |
| Leukocyte infiltration [n (%)] | 16 (100.0%) |
| Degree of leukocyte infiltration [n (%)]: | |
| + | 8 (50.0%) |
| ++ | 8 (50.0%) |
| +++ | 0 (0.0%) |
| Plaques containing macrophages [n (%)] | 12 (75.0%) |
| Number of macrophages per field of view (M [Q1;Q3]) | 3.0 [3.0; 4.0] |
| Plaques containing lymphocytes [n (%)] | 16 (100.0%) |
| Number of lymphocytes per field of view (M [Q1;Q3]) | 9.5 [6.0; 12.3] |
| Plaques containing neutrophils [n (%)] | 8 (50.0%) |
| Number of neutrophils per field of view (M [Q1;Q3]) | 2.0 [1.0; 2.0] |
| Plaques containing foam cells [n (%)] | 5 (31.3%) |
| Number of foam cells per field of view (M [Q1;Q3]) | 5.0 [3.0; 10.0] |
| **Assessment of neovascularization sites** | |
| Plaques with neovascularization sites | 6 (37.5%) |
| Recanalization [n (%)] | 1 (6.3%) |
| Recanalization + endothelium [n (%)] | 1 (6.3%) |

**Table SM2 (continued)**

| Assessment of fibrosis | |
|---|---|
| The degree of the vessel fibrosis [n (%)] | |
| + | 3 (18.8%) |
| ++ | 11 (68.8%) |
| +++ | 2 (12.5%) |
| The degree of the lipid core fibrosis [n (%)] | |
| + | 7 (43.8%) |
| ++ | 7 (43.8%) |
| +++ | 2 (12.5%) |
| The degree of collagen fragmentation [n (%)]: | |
| + | 5 (31.3%) |
| ++ | 6 (37.5%) |
| +++ | 5 (31.3%) |
| AHA plaque type (Stary et al. 1995) [n (%)]: | |
| III, | 1 (6.3%) |
| IV, | 6 (37.5%) |
| IV-V, | 0 (0.0%) |
| V, | 1 (6.3%) |
| including Va, | 1 (6.3%) |
| Vc, | 0 (0.0%) |
| VI, | 8 (50.0%) |
| including VIa, | 1 (6.3%) |
| VIb, | 5 (31.3%) |
| VIc | 2 (12.5%) |

**Table SM3.** Clinical characteristics of participants included in the Infinium MethylationEPIC BeadChip (Illumina) microarray study

| Parameter | Patients with advanced carotid atherosclerosis (n=16) |
|---|---|
| **Clinical parameters** | |
| Men: Women | 10:6 |
| Age, years (M [Q1;Q3]) | 65.0 [62.5; 69.0] |
| BMI, kg/m$^2$ (M [Q1; Q3]) | 28.4 [26.6; 31.1] |
| Metabolic syndrome [n (%)] | 15 (93.8%) |
| Waist circumference, cm (M [Q1;Q3]) | *no data* |
| History of angina [n (%)] | 11 (68.8%) |
| Angina functional class [n (%)]<br>- I FC<br>- II FC<br>- III FC | <br>2 (12.5%)<br>9 (56.3%)<br>0 (0.0%) |
| History of myocardial infarction [n (%)] | 3 (18.8%) |
| History of coronary artery disease [n (%)] | 14 (87.5%) |
| Duration of coronary artery disease, years (M [Q1;Q3]) | 8.0 [5.8; 9.3] |
| History of chronic heart failure [n (%)] | 15 (93.8%) |
| Chronic heart failure functional class [n (%)]<br>- I FC<br>- II FC<br>- III FC | <br>0 (0.0%)<br>13 (81.3%)<br>0 (0.0%) |
| History of TIA or stroke [n (%)] | 5 (31.3%) |
| Hypertension [n (%)] | 16 (100.0%) |
| Duration of hypertension, years (M [Q1;Q3]) | 16.0 [10.0; 23.3] |
| Hypercholesterolemia [n (%)] | 16 (100.0%) |
| Smoking [n (%)]<br>including former smokers | 12 (75.0%)<br>0 (0.0%) |
| Diabetes mellitus [n (%)] | 4 (25.0%) |
| Chronic limb ischemia [n (%)] | 3 (18.8%) |
| **Laboratory data (M [Q1; Q3])** | |
| Glucose, mmol/l | 5.7 [5.3; 7.6] |
| Total cholesterol, mmol/l | *no data* |
| Triglycerides, mmol/l | *no data* |
| LDL, mmol/l | *no data* |
| HDL, mmol/l | *no data* |
| Atherogenic index | *no data* |
| **Medications before hospitalization [n (%)]** | |
| Aspirin | 16 (100.0%) |
| Clopidogrel | 2 (12.5%) |
| Warfarin | 0 (0.0%) |
| Beta-blockers | 15 (93.8%) |
| Angiotensin converting enzyme inhibitors | 9 (56.3%) |
| Angiotensin II receptor blockers | 5 (31.3%) |
| Calcium channel blockers (dihydropyridine) | 11 (68.8%) |
| Calcium channel blockers (non-dihydropyridine) | 0 (0.0%) |
| Digoxin | 0 (0.0%) |
| Loop diuretics | 0 (0.0%) |

**Table SM3 (continued)**

| Medications before hospitalization [n (%)] | |
|---|---|
| Thiazide diuretics | 2 (12.5%) |
| Spironolactone | 1 (6.3%) |
| Nitrates | 0 (0.0%) |
| Statins | 16 (100.0%) |
| Cordarone | 0 (0.0%) |
| Insulin | 0 (0.0%) |
| Metformin | 2 (12.5%) |

#### 3 Selection of miRNA and *MIR* gene regions for DNA methylation analysis.

We reviewed the literature and identified the microRNAs whose expression in the carotid artery tissue is associated with atherosclerotic lesion formation or plaque instability in at least two studies (Table SM4).

**Table SM4**. Expression of microRNAs associated with carotid atherosclerotic plaque instability in humans

| № | miR | Unstable vs Stable Atherosclerotic Plaque | Study | Atherosclerotic plaque vs intact tissues | Study |
|---|---|---|---|---|---|
| **1** | -21 | ↓ S (TIA or stroke ≤6 weeks before CEA) | Markus B. et al., 2016 | ↑ | Raitoharju E. et al., 2011; Cao J. et al., 2014; Markus B. et al., 2016 |
| | | ↓ S (TIA or stroke ≤6 weeks before CEA), MRA (irregularities of plaque surface), H (ruptured) | Wang H. et al., 2019 | | |
| | | ↓ S (stroke or TIA within the past >14 days), H (ruptured) | Jin H. et al., 2018 | | |
| **2** | -100 | ↑ S | Cipollone F. et al., 2011; Maitrias P. et al., 2015 | ND | |
| **3** | -127 | ↑ S | Cipollone F. et al., 2011; Maitrias P. et al., 2015 | ND | |
| **4** | -133a | ↑ S | Cipollone F. et al., 2011; Maitrias P. et al., 2015 | ND | |
| **5** | -133b | ↑ S | Cipollone F. et al., 2011 | ND | |
| | | ↑ H (low-calcified) | Katano H. et al., 2018 | | |
| **6** | -143 | ↓ S (TIA or stroke ≤6 weeks before CEA) | Markus B. et al., 2016 | ↑ | Markus B. et al., 2016 |
| | | ↓ S (TIA or stroke ≤6 weeks before CEA), MRA (irregularities of plaque surface), H (ruptured) | Wang H. et al., 2019 | ↓ | González-López et al., 2022 |
| **7** | -145 | ↑ S | Cipollone F. et al., 2011; Maitrias P. et al., 2015 | ↓ | Lovren F. et al., 2012 |
| **8** | -146a | ND | | ↑ | Raitoharju E. et al., 2011; Cao J. et al., 2014 |
| **9** | -155 | ND | | ↑ | Raitoharju E. et al., 2011; Nazari-Jahantigh M. et al., 2012 |

**Note.** Atherosclerotic plaque instability was defined by symptoms (S), histology (H), or angiography (A). CEA, carotid endarterectomy; MRA, magnetic resonance angiography; ND, no data available; TIA, transient ischemic attack.

In addition, our previous work identified lower methylation of the CpG island located in the *HOXD4* promoter region, overlapping with the *MIR10B* gene body in the cells of coronary and carotid atherosclerotic plaques, and in peripheral blood leukocytes from patients with atherosclerosis, compared with unaffected internal thoracic arteries and great saphenous veins [Nazarenko M.S. et al., 2015]. Accordingly, the genes encoding the nine miRNAs summarized in **Table SM5** (included miRNA host genes, such as *MIR143HG* (*CARMN*) for miR-143/145 cluster), together with *MIR10B*, were considered for further analysis of the methylation level within *MIR* genes regulatory regions.

Unfortunately, primer design and PCR optimization were unsuccessful for several candidate genes. Therefore, this Supplementary Material describe only the target regions and primers for which all experimental stages were completed and methylation data were obtained (**Table SM6**).

**Table SM5.** Candidate genes and regions for the analysis of miRNA regulatory elements

| miRNA | Gene (locus) | | Coordinates* of the miRNA gene/ CpG island in the miRNA gene (number of CpG sites, if available) | Promoter ID and coordinates* according to FANTOM5 | Coordinates* of the region selected for the methylation level analysis | | miRNA location / host gene/ microRNA cluster |
|---|---|---|---|---|---|---|---|
| miR-10b | *MIR10B* | 2q31.1 | chr2:177,015,031-177,015,140 / chr2:177,014,949-177,015,214 (18 CpG sites) | p1@HOXD3 chr2:177,001,291-177,001,315 | *MIR10B* | chr2:177,014,629-177,015,244 | Intergenic / *HOXD* cluster |
| miR-21 | *MIR21* | 17q23.1 | chr17:57,918,627-57,918,698 / chr17:57,916,300-57,916,700 (16 CpG sites) | p1@AK310806 chr17:57,915,282-57,915,332 | *MIR21* putative promoter | chr17:57,915,138-57,916,025 | Intragenic / *VMP1* intron |
| | | | | | *MIR21* CpG island | chr17:57,915,960-57,916,717 | |
| | | | | | *MIR21* gene body | chr17:57,918,142-57,918,840 | |
| miR-100 | *MIR100* | 11q24.1 | chr11:122,022,937-122,023,016 | p2@ENST00000533109 chr11:122,051,328-122,051,348 | chr11:122,024,891-122,025,506 | | Intragenic / *MIR100HG* intron |
| miR-127 | *MIR127* | 14q32.2 | chr14:101,349,316-101,349,412 / chr14:101,349,373-101,349,860 (36 CpG sites) | p1@MEG3 chr14:101,292,449-101,292,462 | chr14:101,349,259-101,349,884 | | Intergenic / *RTL1* imprinted region / miR-433/127 cluster |
| miR-143 | *MIR143* | 5q32 | chr5:148,808,545-148,808,784 | p1@MIR143HG chr5:148,786,424-148,786,453 | chr5:148,786,304-148,786,864 | | Intragenic / noncoding RNA *CARMN* (*MIR143HG*) / miR-143/145 cluster |
| | | | | | chr5:148,808,001-148,808,583 | | |
| miR-145 | *MIR145* | | chr5:148,810,209-148,810,296 | | – | | |

* All coordinates are given according to the GRCh37/hg19 genome assembly

**Table SM6**: Studied regions and primer structure

| **Region** | **Location relative to the gene** | **CpG-region category*** | **Primer sequence** | **Primer annealing temperature** | **Number of CpG sites** | **Amplicon location** (according to GRCh37/hg19 assembly) | **PCR product length (bp)** |
|---|---|---|---|---|---|---|---|
| *MIR10B* | around the gene body | shore + CpG island | F: GTAGTAGGAAAAGGAGTAGATATAGGATATT<br>R: CCTCCTAAAATCCAAAATAATAAAACAAAA | 62 | 23 | chr2:177014629-177015244 | 616 |
| | | | F: ATTTTGGTAGAAGAATGAGGGAATT<br>R: TTCTTTTCAACACCCAAAAAATACT | 62 | 20 | chr2:177014868-177015332 | 465 |
| *MIR21* putative promoter | upstream | open sea | F: TTTTTTGAGAAGAGGGGATAAGT<br>R: TCAAAACAACAACTCATTCTATATAACAAA | 58 | 9 | chr17:57915138-57916025 | 888 |
| *MIR21* CpG island | upstream | CpG island ** | F: TTAAAAATAAGGGTAGAGTTAATGGAA<br>R: TTAATCAACTAAAAACAACAAAAATCC | 60 | 17 | chr17:57915960-57916717 | 758 |
| *MIR21* gene body | around the gene | open sea | F: GGGATTGGTTTATTTGGGGA<br>R: CCTCCATACTACTACATTATAACACAAAAA | 62 | 6 | chr17:57918142-57918840 | 699 |
| *MIR100* E-box | upstream | open sea | F: TTGATAAAGGTATGGGGTTAGAGGTA<br>R: AATTCTTACTTAATAAAACAAACCAAAAT | 60 | 8 | chr11: 122024891-122025506 | 616 |
| *MIR127* | around the gene body | shore + CpG island | F: TTTTGGTATTTATAGGTTTTTAAGG<br>R: AATAAAAATAAACCCCTACCACAAC | 60 | 40 | chr14:101349259-101349884 | 626 |
| *MIR143HG* promoter | upstream | open sea | F: GGGGTAGGTTAAGGTAAGGTAGTTA<br>R: AAACCAACAAAACCATAAAACTAAAAC | 60 | 10 | chr5:148786304-148786864 | 561 |
| *MIR143* gene body | 5' part of the gene | open sea | F: AGTATAAAGGTGGGGTATGGTATTAAA<br>R: ACAAAACAACTTCTCTCTTCCTAAACT | 60 | 5 | chr5:148808001-148808583 | 583 |

Note: *CpG-region categories were defined by CpG density. A **CpG island** is a dense cluster of CpG sites; a **shore** is a region up to 2 kb upstream or downstream from CpG island; and **open sea** is a region contained single CpG sites at a distance exceeding ± 4000 bp from the CpG island.** Classified as open sea according to UCSC browser.

According to the FANTOM5 database, the regulatory elements of the intergenic miRNAs *MIR10B* and *MIR127* are located at a distance from the miRNA gene bodies (~14 thousand bp upstream for *MIR10B* and ~57 thousand bp upstream for *MIR127*) (**Table SM5**). Previous studies have reported that atherosclerosis could alter the DNA methylation levels within regions in close proximity to the genes of these miRNAs [Aavik E. et al., 2015; Lacey M. et al., 2019]. We therefore selected regions covering CpG islands that overlap the *MIR10B* and *MIR127* gene bodies. According to the UCSC Genome Browser (GRCh37/hg19 Assembly) (http://genome-euro.ucsc.edu), the CpG island in the *MIR10B* gene region is 266 bp at length (chr2:177014949-177015214) and contains 18 CpG sites, whereas the CpG island in the *MIR127* gene region is 488 bp at length (chr14:101349373-101349860) and contains 36 CpG sites.

For the intragenic miRNAs – *MIR21*, *MIR143* and *MIR145* – the reported locations of regulatory regions differed among publications and databases. We therefore visualized the reported coordinates in the UCSC Genome Browser and selected genomic regions that encompassed as many candidate regulatory elements as possible.

Most studies place the *MIR21* promoter within chr17:57914850-57915900 (Supplementary Figure 1), whereas the *MIR21* gene body is located at chr17:57918627-57918698. Iorio M.V. et al. (2007) outlined a CpG island 2.5 kb upstream of the *MIR21* gene body, within approximately chr17:57916300-57916700. This 310-bp region met the following CpG island criteria: CpG content >50%, an observed-to-expected CpG ratio >0.6, and length >200 bp [Iorio M.V. et al., 2007]. Nevertheless, applying the exact same criteria, the UCSC Genome Browser does not detect the CpG island 2.5 kb upstream the *MIR21* gene body. MethPrimer (https://www.urogene.org/cgi-bin/methprimer/methprimer.cgi) identified a 132-bp CpG island at chr17:57916335-57916466, with CpG content of 40% and an observed-to-expected CpG ratio >0.6. On this basis, we selected three regions for methylation analysis of the *MIR21* regulatory elements: the putative promoter of the *MIR21* gene, the CpG island and the *MIR21* gene body (**Table SM6**).

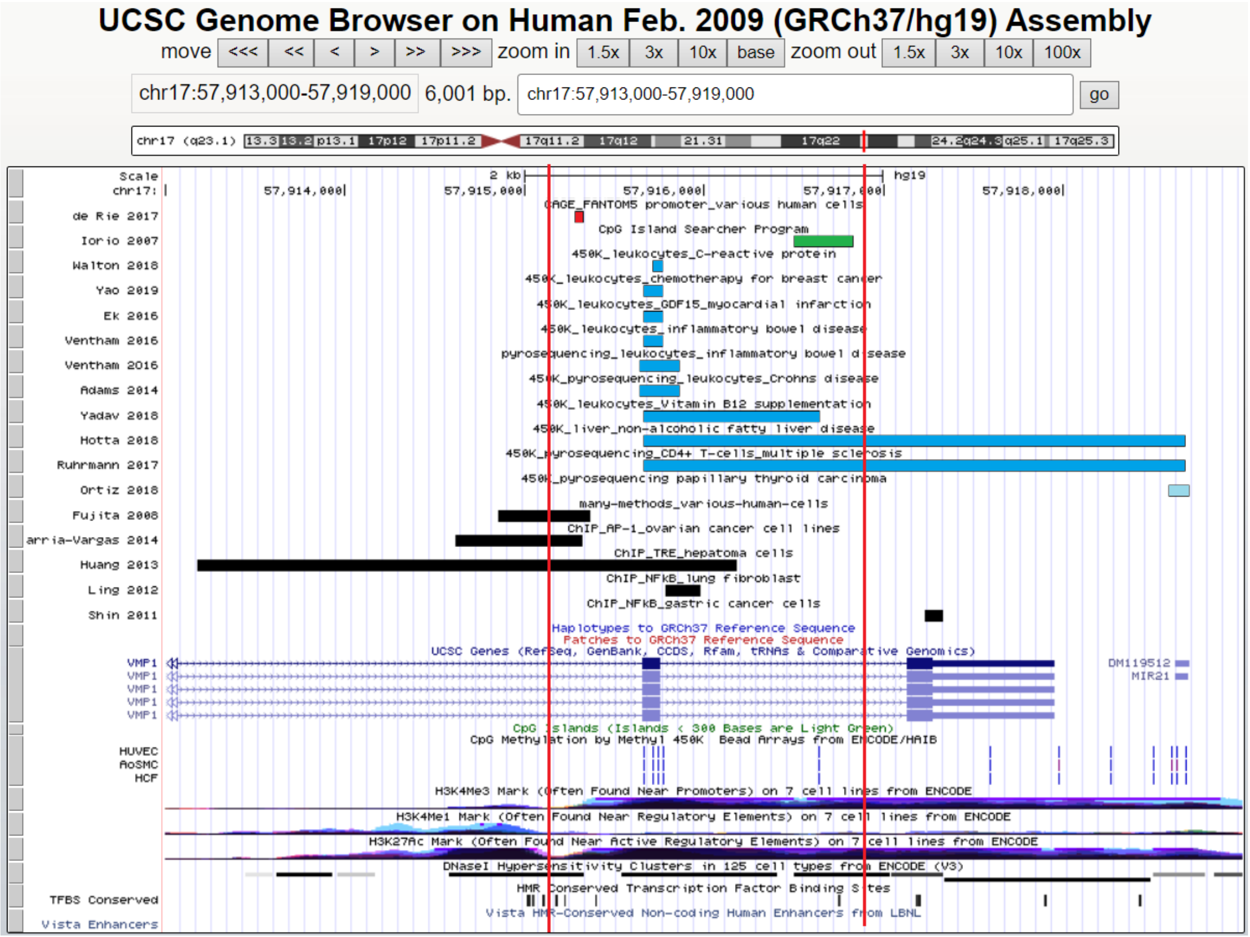

**Supplementary Figure 1.** Determining the location of regulatory elements of the *MIR21* gene.

The red box indicates the promoter region specified in the FANTOM5 database (http://fantom.gsc.riken.jp/5/ [de Rie D. et al., 2017]), green box indicates the CpG island [Iorio M.V. et al., 2007], blue boxes indicate the regions for which the DNA methylation level was studied using microarray methods and pyrosequencing in blood leukocytes and liver tissue in various diseases [Adams A.T. et al., 2014; Ek W.E. et al., 2016; Ventham N.T. et al., 2016; Hotta K. et al., 2018; Ruhrmann S., et al., 2018; Walton E. et al., 2018; Yadav D.K et al., 2018; Yao S. et al., 2019], light blue box indicates the region that was demethylated after 5-azacytidine treatment of papillary thyroid carcinoma cells [Ortiz I.M.D.P. et al., 2018], black boxes indicate the putative promoters based on the computational approach [Fujita S. et al., 2008] and the chromatin-immunoprecipitation sequencing (ChIP-seq) data from the transcription factors search [Shin V.Y. et al., 2010; Ling M. et al., 2012; Huang Y.H. et al., 2013; Echevarría-Vargas I.M. et al., 2014]. The vertical lines delimit the region selected for analysis.

Information about the own promoter of the *MIR143* gene is limited. The *MIR143* gene body is located at chr5:148808545-148808784. A 26-kb aorta/ovary/heart super-enhancer spanning the *CARMN* (*MIR143HG*) region has been reported to upregulate *MIR143* and *MIR145* irrespective of the precise transcription start sites of these miRNA genes and may be susceptible to disease-associated downregulation through DNA hypermethylation [Ehrlich K.C.et al., 2019. Other studies indicate that miR-143 expression may be regulated by the *MIR143HG* (miR-143 host gene) promoter at chr5:148786424-148786453 (**Supplementary Figure 2**) [Blick C. et al., 2015; Deng L.et al., 2015; de Rie D. et al., 2017; Riches K. et al., 2017]. We therefore selected the *MIR143HG* (*CARMN*) promoter region at chr5:148786304-148786864 (561 bp, 10 CpG sites) and the *MIR143* gene body region at chr5:148808001-148808583 (583 bp, 5 CpG sites) for methylation analysis.

For *MIR145*, the region selected to encompass as many putative promoter elements as possible (chr5:148810000-148810500) also overlapped the *MIR145* gene body (chr5:148810209-148810296) (**Supplementary Figure 2**). Unfortunately, the of the *MIR145* gene body region (chr5:148809848-148810488) was unsuccessful.

No promoter location had been reported for the intragenic miRNA gene *MIR100*. Chen et al. (2014) showed that transcription factor *ZEB1* binds an E-box element that enhances miR-100 expression. This six-nucleotide motif (CAGCTG) is located 2.2 kb upstream of *MIR100* [Chen D. et al., 2014]. According to the UCSC Genome Browser, the *MIR100* gene body spans chr11:122022937-122023016, and the E-box is located at chr11:122025183-122025188. We therefore selected chr11:122024891-122025506 (616 bp, 8 CpG sites), which fully encompasses the E-box, for methylation analysis. We also selected chr11:122022733-122023292 (560 bp, 7 CpG sites), which overlaps the *MIR100* gene body, but amplification of this region was not effective to obtain a product suitable for sequencing.

In total, eight regions were selected for DNA methylation analysis by BSAS (**Table SM6**): the CpG island overlapping the *MIR10B* gene body; the putative promoter, CpG island, and gene body of *MIR21*; the *MIR100* E-box region containing the *ZEB1*-binding site; the CpG island overlapping the *MIR127* gene body; the *MIR143HG* promoter; and the *MIR143* gene body.

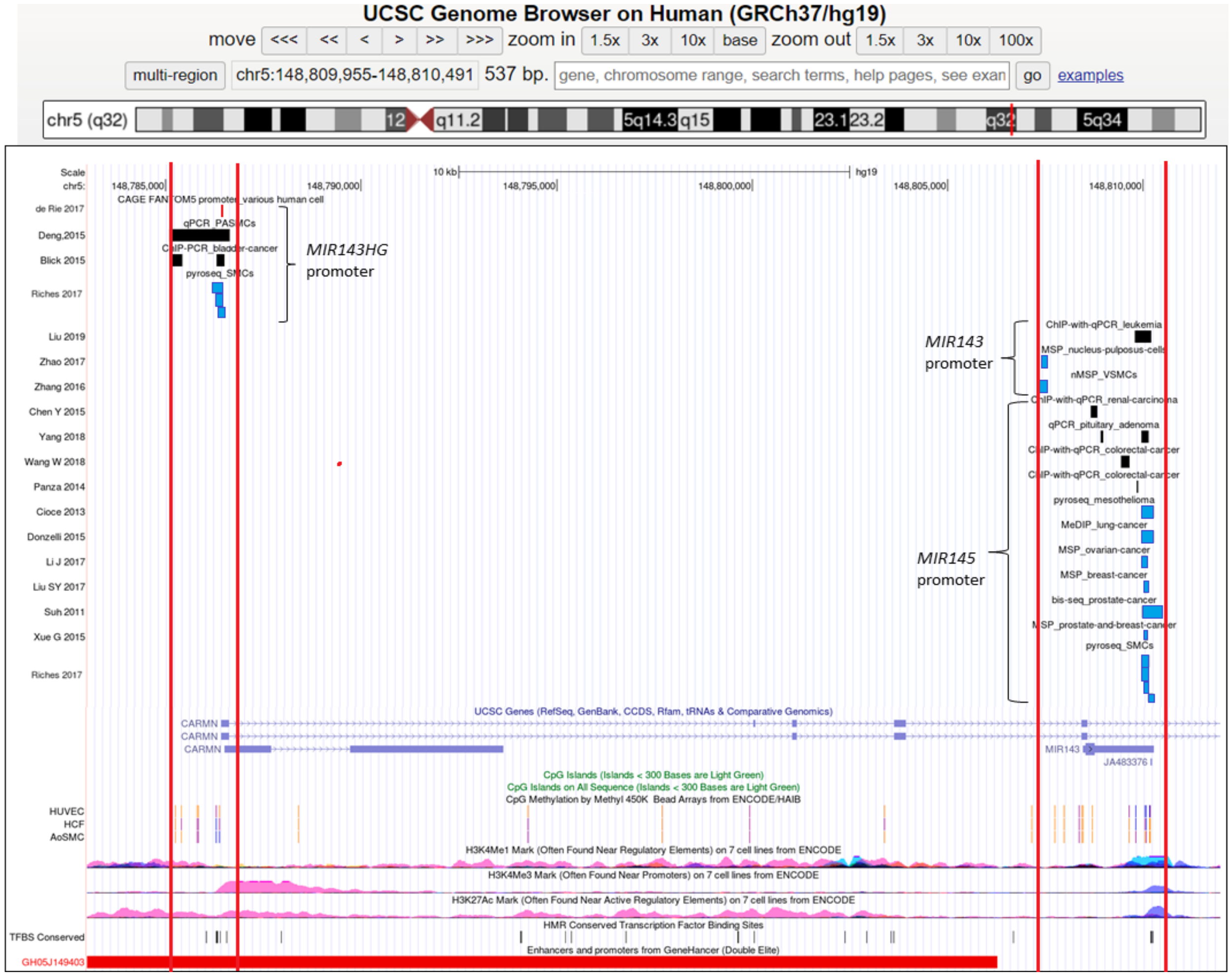


**Supplementary Figure 2. Determining the location of regulatory elements of the *MIR143* and *MIR145* genes.**

The red box indicates the promoter region specified in the FANTOM5 database (http://fantom.gsc.riken.jp/5/ ) [de Rie D. et al., 2017], blue boxes indicate the regions previously examined for DNA methylation in smooth muscle cells [Zhang H.P. et al., 2016; Riches K. et al., 2017], nucleus pulposus cells [Zhao K.et al., 2017] and in various cancer cells [Suh S.O. et al., 2011; Cioce M. et al., 2013; Donzelli S. et al., 2015; Xue et al., 2015; Li J.et al., 2017; Liu J. et al., 2017; Liu S.Y. et al., 2017], black boxes indicate the regions proposed as *MIR143* or *MIR145* promoters, including findings based on the chromatin-immunoprecipitation sequencing (ChIP-seq) data from the transcription factors search [Panza A. et al., 2014; Blick C. et al., 2015; Chen Y. et al., 2015; Deng L.et al., 2015; Wang W. et al., 2018; Yang W. et al., 2018; Liu T. et al., 2019]. The vertical lines delimit the region selected for analysis.

#### 4 DNA methylation analysis by Bisulfite Amplicon Sequencing (BSAS)

DNA methylation analysis by high-throughput MPS included the following steps: (1) selection of target genomic regions and primer design; (2) amplification of target regions by PCR; (3) equimolar pooling and purification of amplicons; (4) DNA library preparation and sequencing; (5) bioinformatics processing of the results.

**4.1 Primer design for BSAS.** Primers were designed several publicly available bioinformatic tools. MethPrimer (https://www.methprimer.com/) was used with the following criteria: primer length, 20-30 nucleotides; melting temperature, 58-65°C; and product size, 300-900 nucleotides [Li L.C., Dahiya R., 2002]. The theoretical melting temperature and GC content of the oligonucleotide sequence, as well as the possibility of hairpins, self- and hetero-dimers of primers were evaluated using the IDT OligoAnalyzer online tool (https://eu.idtdna.com/calc/analyzer). Amplicon specificity for the target region was determined by *in silico* PCR using BiSearch ePCR online tool (http://bisearch.enzim.hu/?m=genompsearch). Primer sequences are provided in in **Table SM6**.

**4.2. Amplification of target DNA regions.** Target regions were amplified using the BioMaster HS-Taq PCR-Color (2×) PCR kit (BioLabMix, Russia). The optimal annealing temperature was determined by gradient PCR, varying the temperature within ±2°C of the calculated melting temperature for each primer pair. DNA amplification was performed on ProFlex PCR System (ThermoFisher Scientific, USA) and DTPrime4 (DNA-Technology, Russia) using the following program: 95°C for 5 min, 40 cycles of 95°C for 30 s, primer-specific annealing temperature for 30 s, and 72°C for 60 s; followed by 72°C for 5 min. Annealing temperatures for individual primer pairs are listed in **Table SM6**. The PCR products were evaluated by DNA electrophoresis in 3% agarose gel stained with ethidium bromide and visualized using a ChemiDoc XRS+ System or Gel Doc XR+ Imaging System (Bio-Rad, USA).

**4.3 Equimolar pooling and purification of amplicons.** Amplicons from each DNA sample were pooled equimolarly (approximately 200 billion copies of double-stranded DNA per amplicon) and diluted in 200 uL. Pooled amplicons were purified for library preparation using the GeneJET NGS PCR Product Cleanup Kit (ThermoFisher Scientific, USA). The concentration of purified amplicons was measured on a Qubit 3.0 fluorimeter (ThermoFisher Scientific, USA).

**4.4. DNA library preparation and sequencing.** After DNA concentration measurement, purified amplicon pools were used to prepare DNA libraries with the Nextera XT DNA Library Prep Kit or Nextera DNA Flex Library Prep Kit (Illumina, USA) according to the manufacturer's protocol. Pooled libraries were sequenced on a MiSeq (Illumina, USA) using MiSeq Reagent Kit v2 (300-cycles, Illumina, USA) in the sequencing mode of 150-bp paired-end reads.

**4.5. Bioinformatic processing of the results.** The *methylseq* pipeline implemented in Nextflow [Ewels P.A. et al., 2020] was used to automate and standardize data processing. The resulting reads of DNA fragments were automatically sorted into samples with the nucleotide sequence of the adapters removed. Reads were filtered for quality (minimum Q30) and length (minimum 20 nucleotides) using the *Trim Galore* tool [Krueger F. et al., 2021], then aligned to the GRCh37/hg19 human reference genome using the *bwa-meth* tool [Pedersen B.S. et al., 2014]. CpG methylation information was extracted from BAM files using Bismark tool [Krueger F., Andrews S.R., 2011]. FastQC (https://www.bioinformatics.babraham.ac.uk/projects/fastqc/), Qualimap, Preseq, and MultiQC tools were used for quality control [Daley T., Smith A.D., 2013; Ewels P. et al., 2016; Okonechnikov K. et al., 2016].

Subsequent processing of locus-specific methylation profiles and statistical analysis were performed in R using methylKit [Akalin A. et al., 2012). CpG-site methylation was defined as the percentage of reads containing methylated cytosine relative to all cytosine reads at the corresponding genomic position. CpG sites with less than 15-fold coverage were excluded from the analysis.
